# Joint associations between objectively measured physical activity volume and intensity with body-fatness. The Fenland Study

**DOI:** 10.1101/2021.03.26.21254409

**Authors:** Tim Lindsay, Katrien Wijndaele, Kate Westgate, Paddy Dempsey, Tessa Strain, Emanuella De Lucia Rolfe, Nita G Forouhi, Simon Griffin, Nick J Wareham, Søren Brage

## Abstract

**Background/Objectives:** Physical activity energy expenditure (PAEE) represents the total volume of all physical activity. This can be accumulated as different underlying intensity profiles. Although volume and intensity have been studied in isolation, less is known about their joint association with health. We examined this association with body-fatness in a population-based sample of middle-aged British women and men.

**Methods:** 6148 women and 5320 men from the Fenland study with objectively-measured physical activity from individually calibrated combined heart rate and movement sensing and DXA-derived body-fat percentage (BF%) were included in the analyses. We used linear and compositional isocaloric substitution analysis to examine associations of PAEE and its intensity composition with body-fatness. Sex-stratified models were adjusted for socio-economic and dietary covariates.

**Results:** PAEE was inversely associated with body-fatness in women (beta=-0.16 (95%CI: −0.17; −0.15) BF% per kJ·day^-1^·kg^-1^) and men (beta=−0.09 (95%CI: −0.10; −0.08) BF% per kJ·day^-1^·kg^-1^). Intensity composition was significantly associated with body-fatness, beyond that of PAEE; the reallocation of energy to vigorous physical activity (>6 METs) from other intensities was associated with less body-fatness, whereas light activity (1.5-3 METs) was positively associated. However, light activity was the main driver of overall PAEE volume, and the relative importance of intensity was marginal compared to that of volume; the difference between PAEE in tertile 1 and 2 in women was associated with 3 percentage-point lower BF%. Higher vigorous physical activity in the same group to the maximum observed value was associated with 1 percentage-point lower BF%.

**Conclusions:** In this large, population-based cohort study with objective measures, PAEE was inversely associated with body-fatness. Beyond the PAEE association, greater levels of intense activity were also associated with lower body-fatness. This contribution was marginal relative to PAEE. These findings support current guidelines for obesity prevention which emphasise moving more over the specific intensity or duration of that activity.

## Introduction

Physical activity energy expenditure (PAEE) is the most variable component of total energy expenditure (TEE) and is within voluntary control in free-living settings. Physical activity is also characterised by its intensity, which is usually expressed as the metabolic equivalent of task (MET), with 1 MET corresponding to resting metabolic rate. As energy expenditure is a function of intensity and time, it follows that many different behavioural patterns, or intensity profiles, can underpin any given total volume of PAEE. For example, large amounts of time spent in light physical activity (LPA), or a lesser amount of time spent in moderate physical activity (MPA), or an even shorter period of time spent in vigorous physical activity (VPA) could all lead to the same PAEE.

Several studies have shown that physical activity is associated with a range of metabolic conditions, including obesity or hard end-points such as mortality^1–5^. Typically, overall volume and intensity of physical activity (PA) have been examined in parallel, and less is known about the role of intensity beyond that of volume. In order to advance this understanding, these two dimensions of physical activity need to be examined in an integrated model.

Recently, a number of isotemporal substitution studies have sought to examine the associations between the reallocation of time in one type of behaviour for another. For example, time spent sedentary for time in moderate-to-vigorous physical activity (MVPA)^4, 6–8^. However, such models do not account for the inherent increase in PAEE that occurs when a fixed amount of time is reallocated to a more intense behaviour. Simultaneous examination of volume and intensity therefore requires a methodological adaptation to enable integrated analysis.

Examples of comparable integrated analyses exist in nutritional epidemiology, where the combined role of total energy intake and macronutrient energy composition are studied simultaneously in the general population^9, 10^. In essence, these studies evaluate whether it is the total number of calories, or their source or exchanges between the different sources, which drives associations with health.

In contrast, little is known about the role of the energy expenditure composition of physical activity in determining body-fatness, beyond that of total PAEE. Investigating this in free-living individuals requires valid energy expenditure estimates across the full spectrum of intensity and well-characterised body-fatness in population studies, large enough that different combinations of high and low PA volumes with different intensity profiles naturally occur. Here, we aimed to evaluate the integrated joint associations between objectively measured PA volume, intensity and body-fatness in a large sample of free-living adults.

## Methods

### Participants

The Fenland Study is an ongoing population-based observational study of 12 435 young and middle-aged adults, the research methods for which have been previously described (DOI: https://doi.org/10.1186/ISRCTN72077169)^11^.

Briefly, participants born between 1950 and 1975 were recruited between 2005 and 2015 from general practice lists in the East of England, UK. Exclusion criteria were pregnancy, physician-diagnosed diabetes, inability to walk unaided, psychosis, and terminal illness. All participants provided written informed consent and the study was approved by the local ethics committee (NRES Committee – East of England Cambridge Central) and performed in accordance with the Declaration of Helsinki.

### Measures

#### Anthropometry and other clinical measures

Participants attended a clinical research facility after an overnight fast. All measurements were taken by trained research staff following standardised procedures. Height (cm) was measured with a rigid, wall mounted stadiometer (SECA 240; Seca, Birmingham, UK) and weight (kg) was measured in light clothing with calibrated scales (TANITA model BC-418 MA; Tanita, Tokyo, Japan).

Total body fat mass (FM, in grams) was determined by dual-energy X-ray absorptiometry using a Lunar Prodigy Advanced with the enCORE™ software version 14.10.022 (GE Medical Healthcare, Hatfield, UK) as validated against the gold-standard 4-compartment method^12^. Standard imaging and positioning protocols were applied. Briefly, the system was first calibrated using a spine phantom made of calcium hydroxyapatite, embedded in a lucite block. The coefficient of variation for scanning precision was 2% for total fat mass (30 consecutive scans). To scan participants, they were positioned lying supine within the scanning area of the DXA bed. Anatomical regional boundaries were demarcated and corrected if necessary. For the present analysis, we excluded those with missing DXA data (n=566) but included DXA-scanned participants with medical implants, amputees and minor scanning artefacts (n=135). For participants who were too large to fit within the scanning area (n=108), the symmetry method was used during image processing, e.g. an unscanned left arm was assumed to match the right arm. The primary outcome variable was body fat percentage (BF%) and the secondary outcome was fat mass index (FMI, calculated as fat mass divided by height squared).

#### Physical activity assessment

Physical activity was measured objectively by fitting participants with a combined heart rate and uniaxial movement sensor (Actiheart, CamNtech, Papworth, UK), attached to the chest with standard ECG electrodes^13^. Heart rate was individually calibrated using a treadmill test as previously described^14^. At the end of the clinical visit, participants were asked to wear the sensor, initialised to collect data at 1-min resolution, for the following 6 days, and to return the monitor by freepost. Participants were advised that the device was waterproof and should be worn continuously, including during showering, water-based activities, and sleeping, whilst continuing with their usual activities. It could be removed to change electrodes, spare sets of which were provided.

Heart rate data were pre-processed^15^, individually calibrated^14^, and combined with acceleration to estimate instantaneous PAEE (intensity) according to methods previously described and validated^16, 17^. Energy spent at each PA intensity level was calculated as a fraction of PAEE by dividing energy expended at that level by total PAEE. We did this for multiple intensity categories in high resolution (every 0.25 MET) and also grouped into the broader categories of sedentary behaviour/sleep (SS: <1.5 METs), light physical activity (LPA: 1.5-3 METs), moderate physical activity (MPA: 3-6 METs), and vigorous physical activity (VPA: >6 METs), where 1 MET = 71 J·min^-1^·kg^-1^ (∼3.5 ml O_2_·min^-1^·kg^-1^). As a sensitivity analysis, we redefined the non-sedentary categories as LPA (1.5-4 METs), MPA (4-7 METs), and VPA (>7 METs). For individuals registering no MPA or VPA time, these analytical fractions were replaced with a value of 0.0001 below the lowest recorded non-zero value for the population, since some analyses require log-transformation of these exposure variables.

Participants were excluded from the present analysis if they had failed to wear their sensor for at least 72 hours overall (n=319), or at least 8 hours cumulative wear for each quadrant of the day (3am to 9am, 9am to 3pm, 3pm to 9pm, 9pm to 3am); these latter criteria ensure behavioural information is available for all parts of the day from at least two different days. Furthermore, activity records were excluded if they did not measure 0 m·s^-2^ (zero movement) at some point during the monitoring period as this indicates a technical problem with the acceleration sensor (n=76).

#### Covariates

Demographic, lifestyle and health variables were collected using self-report. These included age, sex, marital status (single, married, widowed/separated/divorced), education (compulsory, further – A-level/apprenticeship/sub-degree level, higher – degree level or above), household income level (<£20,000, £20,000-£40,000, >£40,000), ethnicity (White, South Asian, Black, East Asian, Others and unknown), location (Cambridge, Ely, Wisbech), and smoking status (never, ex-smoker, current). Participants with missing socio-demographic covariate data were retained for analysis; missing data in categorical variables were coded as a separate category. Habitual diet over the previous year was self-reported using a validated food frequency questionnaire (FFQ)^18^, from which estimated total energy intake (kJ·day^-1^) from carbohydrates, protein, fats, and alcohol were derived. Participants with no dietary information were excluded from the analysis (n=6). Plasma vitamin C was measured as a biomarker of fruit and vegetable intake and indicator of overall diet quality^19^. Participants with plasma vitamin C levels below the assay detection threshold were coded at the minimum detectable level (n=45). Participants missing this measure were included in analysis by imputation from age, sex, self-reported fruit and vegetable intake and vitamin supplementation (n=263).

### Data analysis

All analyses were stratified by sex owing to fundamentally different adiposity patterns between men and women. For sample characteristics, means and standard deviations are reported for continuous variables and proportions are reported for categorical variables. All analyses were undertaken in Stata 14 (StataCorp, Texas).

#### Associations between PAEE and intensity of PA on body-fatness

Multivariable linear regression was used to examine the associations between body-fatness outcomes (BF% and FMI) and the PA exposure variables, with all models controlling for the potential confounding variables described above.

First, we examined the association between total PAEE as a single exposure and both adiposity outcomes. We then explored the relative association of PA intensity through a series of sequential linear regressions for the fraction of PAEE spent above a given intensity threshold (1-11 METs), controlling for overall PAEE. Beta coefficients were graphically represented to estimate the difference in BF% associated with a 1% difference in PAEE above a given MET threshold.

#### Isocaloric substitution analysis

To assess the combined association between the relative PA intensities and body-fatness, we simulated the effect of reallocating energy expenditure between the four broader intensity categories on body adiposity, using two types of models. First, we used linear isocaloric substitution analysis as per methods previously described^20^, except with percentage of PAEE substituted, rather than time. In brief, each iteration of the model simultaneously included total PAEE and its fractions expended in the intensity categories, systematically excluding (dropping) the fraction of energy expended in one intensity category. Regression coefficients (95% CI) of the intensity components included in these models provide an estimate of the change in the outcome variable when reallocating energy expended in the dropped intensity to another by percentage of total PAEE, while decreasing energy expended in the dropped intensity category by the same percentage. We checked appropriateness of the linear approximation of these relationships by visual inspection of scatterplots for each exposure-outcome, controlled (residualised) for the other covariates in the model.

As an alternative approach to assess the combined association between the relative PA intensities and body-fatness, we modelled the composition of PAEE using a compositional data analysis approach according to methods previously described^4, 7^. Fractional energy spent at each intensity of PA was transformed into sets of isometric log-ratios (ILRs). Each set of ILRs was regressed against the outcome of interest, controlling for total PAEE and confounders to produce a set of beta coefficients. These beta coefficients were then exponentiated (back transformed) and scaled with the geometric mean of PAEE in the relevant energy intensity for the population stratum of interest (men, women, tertile of PAEE). These results were then plotted against reallocations of PAEE accumulated at the given intensity in kJ·day^-1^·kg^-1^, whilst holding total PAEE constant. To provide an equivalent of linear isocaloric substitution using compositional analysis, pairwise reallocations were also modelled^6^. For all models, variance inflation factors were checked to assess potential issues of collinearity, and distribution of residuals were visually inspected for approximation to normality.

## Results

### Participant characteristics

A total of 11 468 participants (6148 women, 5320 men) were included in the analyses (Table 1). Overall, women and men were of similar age, but women had lower body mass index (BMI) and higher FMI and BF% than men.

**Table 1:**
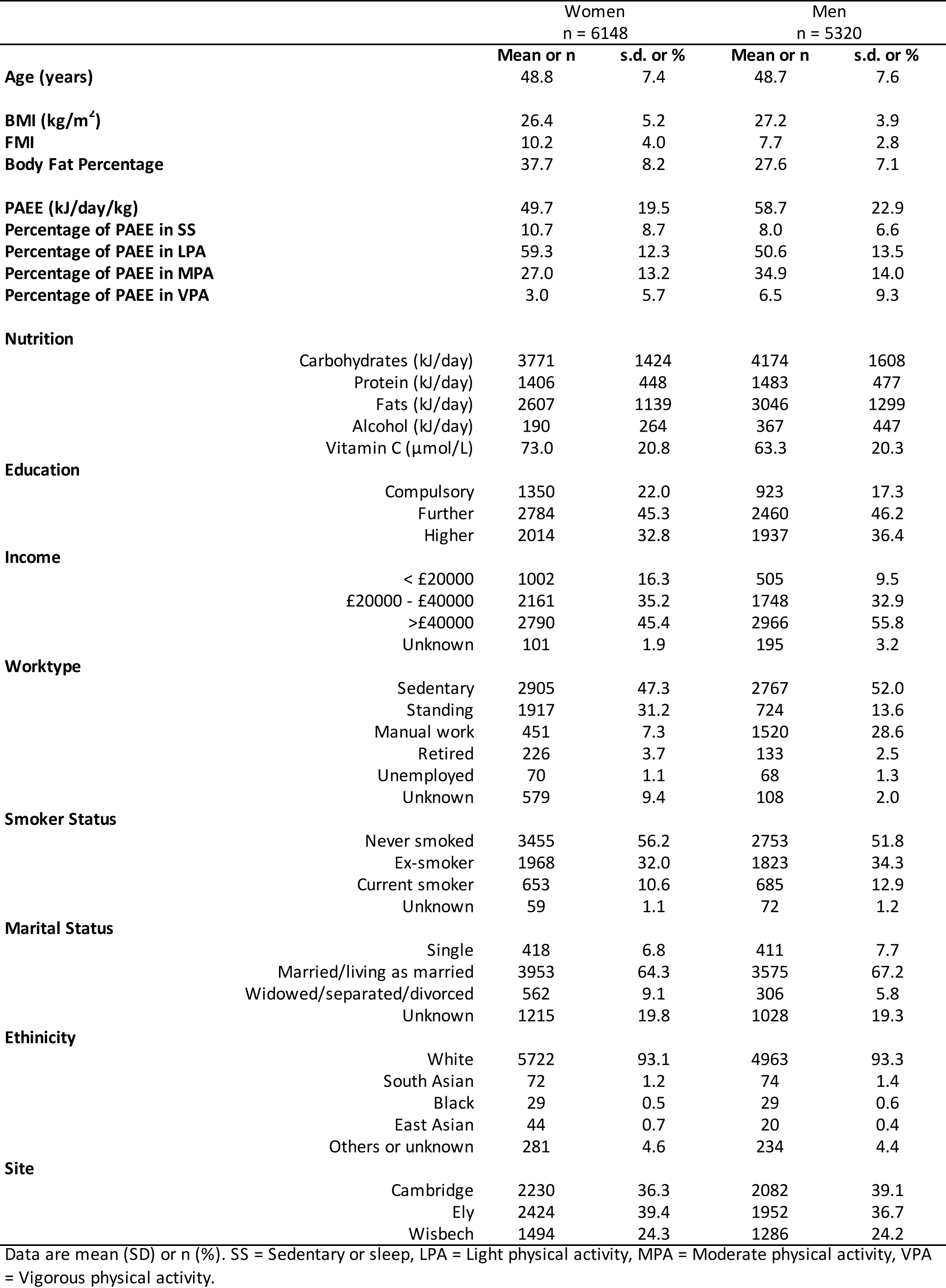
Participant characteristics. The Fenland study 2005-2015.

Women also accumulated lower levels of total PAEE, with a mean (SD) of 50 (20) compared to 59 (23) kJ·day^-1^·kg^-1^ in men. Women and men accumulated PAEE differently, with LPA, MPA, and VPA respectively constituting 59%, 27%, and 3% of women’s total PAEE on average; the comparable figures for men were 51%, 35%, and 7%.

### Associations between PAEE and intensity of PA on body-fatness

Higher levels of PAEE were linearly associated with lower BF% and FMI in both sexes (Figure 1); the adjusted beta coefficients (95%CI) being −0.16 (−0.17; −0.15) %BF per kJ·day^-1^·kg^-1^ in women and −0.09 (−0.10; −0.08) %BF per kJ·day^-1^·kg^-1^ in men. For FMI, corresponding beta coefficients were −0.07 (−0.08; −0.07) kg·m^-2^ per kJ·day^-1^·kg^-1^ in women and −0.03 (−0.04; −0.03) kg·m^-2^ per kJ·day^-1^·kg^-1^ in men.

**Figure 1:**
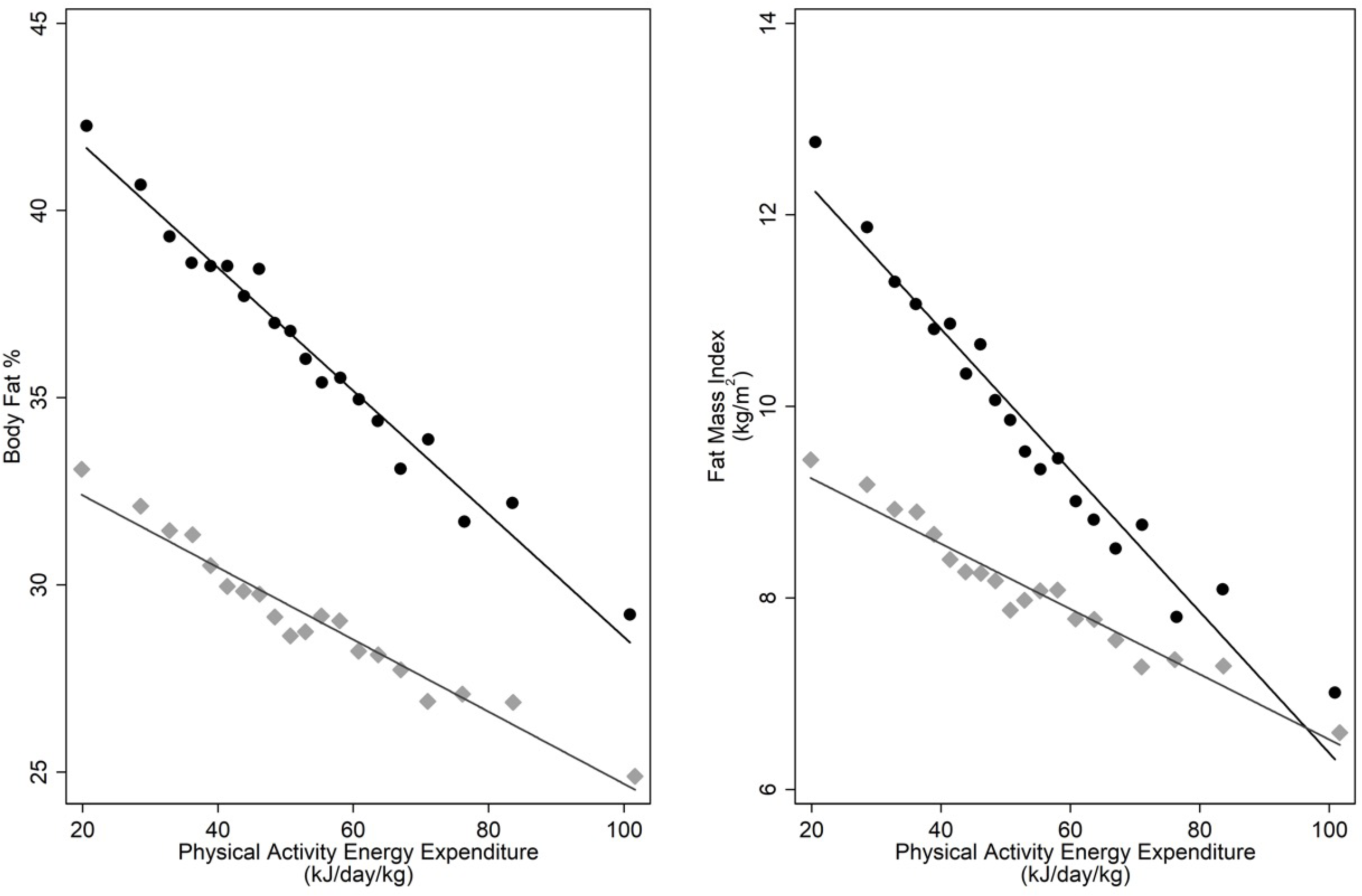
Sex-stratified binscatter of PAEE (kJ/day/kg) and body fat percentage and fat mass index, adjusted for all socio-demographic and dietary covariates. Each bin represents the mean values of 5% of the sex-stratified cohort. Women = grey diamonds, men = black circles.

The relative intensity composition of the accumulated PAEE was also a significant factor in the association with body-fatness. Figure 2 shows the linear beta coefficients for the association with fraction of PAEE accumulated above increasingly higher intensity thresholds, whilst controlling for total PAEE; the association was steeper at higher intensities as indicated by progressively larger negative beta coefficients.

**Figure 2:**
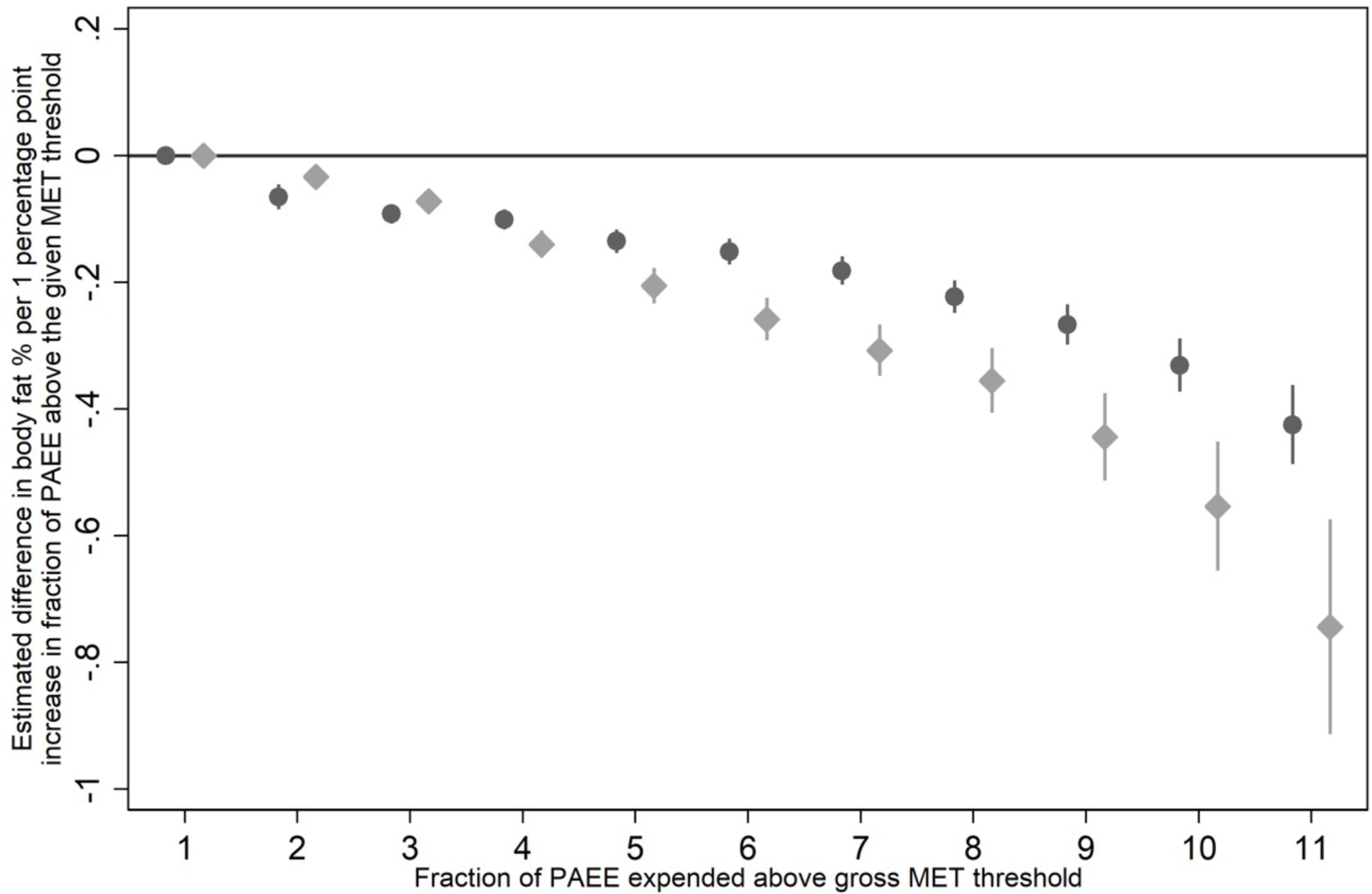
Sex-stratified plot of the beta coefficients from 11 separate, sequential, multivariable linear regressions of the fraction of PAEE spent above each intensity (MET) threshold. Women = light grey diamonds, men = dark grey circles. Error bars are 95% confidence intervals.

### Isocaloric substitution analysis

Table 2 shows the linear isocaloric substitution analyses for the percentage of energy expended at SS, LPA, MPA, or VPA, stratified by sex and adjusted for all dietary and socio-demographic covariates. These integrated analyses model the effect of overall PAEE on body-fatness, while simultaneously simulating the effect of substituting PAEE spent in any one intensity for another. In both sexes, higher levels of PAEE were associated with lower BF%, independent of the underlying intensity distribution. This association was stronger in women than men; each additional 1 kJ·day^-1^·kg^-1^ of active energy expended was associated with a 0.12 percentage-points lower body fat in women, compared to 0.05 percentage-points in men. For a given PAEE volume, the isocaloric substitutions of LPA for energy expended in MPA or VPA, or MPA for energy expended in VPA, were associated with a significantly lower body-fatness. The estimated association was stronger in women than in men (Women: LPA to VPA −0.26 %BF per 1 percentage-point PAEE, MPA to VPA −0.24 %BF per 1 percentage-point PAEE; Men: LPA to VPA −0.17 %BF per 1 percentage-point PAEE, MPA to VPA −0.12 %BF per 1 percentage-point PAEE). These results were not materially different when modelling FMI instead of BF% (Supplementary Table 1). Redefining the intensity categories to LPA (1.5 – 4 METs), MPA (4 – 7 METs), and VPA (>7 METs), produced statistically similar results, with a trend towards increased beta-coefficients for LPA to VPA substitutions (Supplementary Table 2).

**Table 2:**
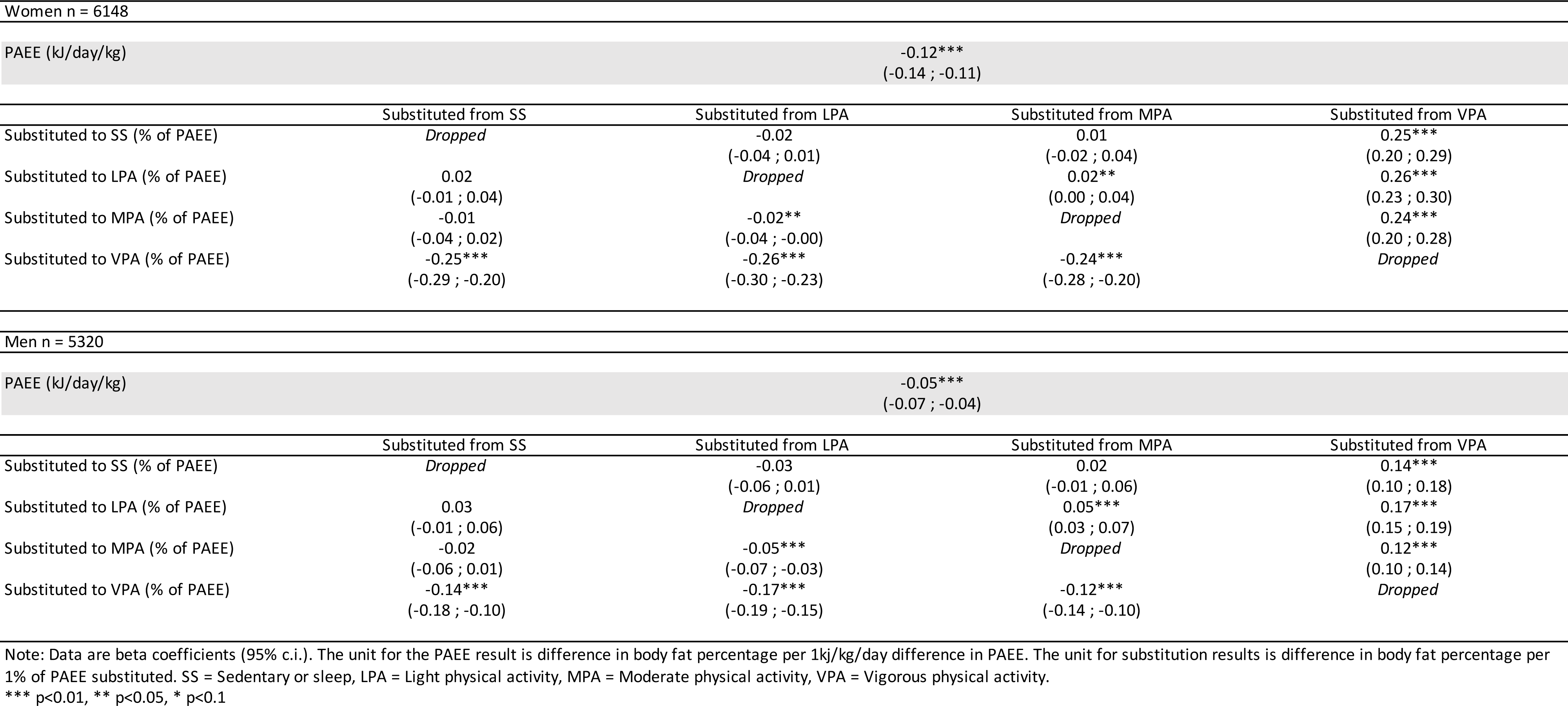
Isocaloric substitution of physical activity energy expenditure and body fat percentage.

Women n = 6148
|  |  |  |  |  |
| --- | --- | --- | --- | --- |
| PAEE (kJ/day/kg) | -0.12***<br>(-0.14 ; -0.11) |  |  |  |
|  | Substituted from SS | Substituted from LPA | Substituted from MPA | Substituted from VPA |
| Substituted to SS (% of PAEE) | *Dropped* | -0.02<br>(-0.04 ; 0.01) | 0.01<br>(-0.02 ; 0.04) | 0.25***<br>(0.20 ; 0.29) |
| Substituted to LPA (% of PAEE) | 0.02<br>(-0.01 ; 0.04) | *Dropped* | 0.02**<br>(0.00 ; 0.04) | 0.26***<br>(0.23 ; 0.30) |
| Substituted to MPA (% of PAEE) | -0.01<br>(-0.04 ; 0.02) | -0.02**<br>(-0.04 ; -0.00) | *Dropped* | 0.24***<br>(0.20 ; 0.28) |
| Substituted to VPA (% of PAEE) | -0.25***<br>(-0.29 ; -0.20) | -0.26***<br>(-0.30 ; -0.23) | -0.24***<br>(-0.28 ; -0.20) | *Dropped* |
| Men n = 5320 |  |  |  |  |
| PAEE (kJ/day/kg) | -0.05***<br>(-0.07 ; -0.04) |  |  |  |
|  | Substituted from SS | Substituted from LPA | Substituted from MPA | Substituted from VPA |
| Substituted to SS (% of PAEE) | *Dropped* | -0.03<br>(-0.06 ; 0.01) | 0.02<br>(-0.01 ; 0.06) | 0.14***<br>(0.10 ; 0.18) |
| Substituted to LPA (% of PAEE) | 0.03<br>(-0.01 ; 0.06) | *Dropped* | 0.05***<br>(0.03 ; 0.07) | 0.17***<br>(0.15 ; 0.19) |
| Substituted to MPA (% of PAEE) | -0.02<br>(-0.06 ; 0.01) | -0.05***<br>(-0.07 ; -0.03) | *Dropped* | 0.12***<br>(0.10 ; 0.14) |
| Substituted to VPA (% of PAEE) | -0.14***<br>(-0.18 ; -0.10) | -0.17***<br>(-0.19 ; -0.15) | -0.12***<br>(-0.14 ; -0.10) | *Dropped* |

Note: Data are beta coefficients (95% c.i.). The unit for the PAEE result is difference in body fat percentage per 1kJ/kg/day difference in PAEE. The unit for substitution results is difference in body fat percentage per 1% of PAEE substituted. SS = Sedentary or sleep, LPA = Light physical activity, MPA = Moderate physical activity, VPA = Vigorous physical activity.
\*\*\* p<0.01, \*\* p<0.05, \* p<0.1

To further examine the relative contribution of intensity and volume, we stratified analyses by PAEE tertile. This stratification revealed a larger linear PAEE beta-coefficient in first tertile women compared to second and third (−0.27, −0.11, and −0.12). Universally across all PAEE tertiles, reallocation of energy to VPA from any other intensity was associated with a significantly lower body fat percentage (Supplementary Table 3).

Using compositional data analysis of the integrated volume-intensity association with body-fatness, overall PAEE remained significantly inversely associated with fatness in both women and men (Supplementary Table 4). The four compositional intensity reallocation curves for each sex are shown in Figure 3, with the accompanying boxplots indicating the relative size of the respective energy reservoirs. These curves show men and women had a similar estimated difference in BF% resulting from an isocaloric 1 kJ·day^-1^·kg^-1^ reallocation of active energy to a specific activity intensity, drawing proportionately from the three other intensities in the composition. Reallocation to VPA was significantly associated with lower body-fatness, whereas reallocation to LPA was significantly associated with higher body-fatness, and reallocation to MPA was not significant. However, reallocation to SS differed between men and women; in men, it was associated with lower body-fatness, whereas in women this association was non-significant.

**Figure 3:**
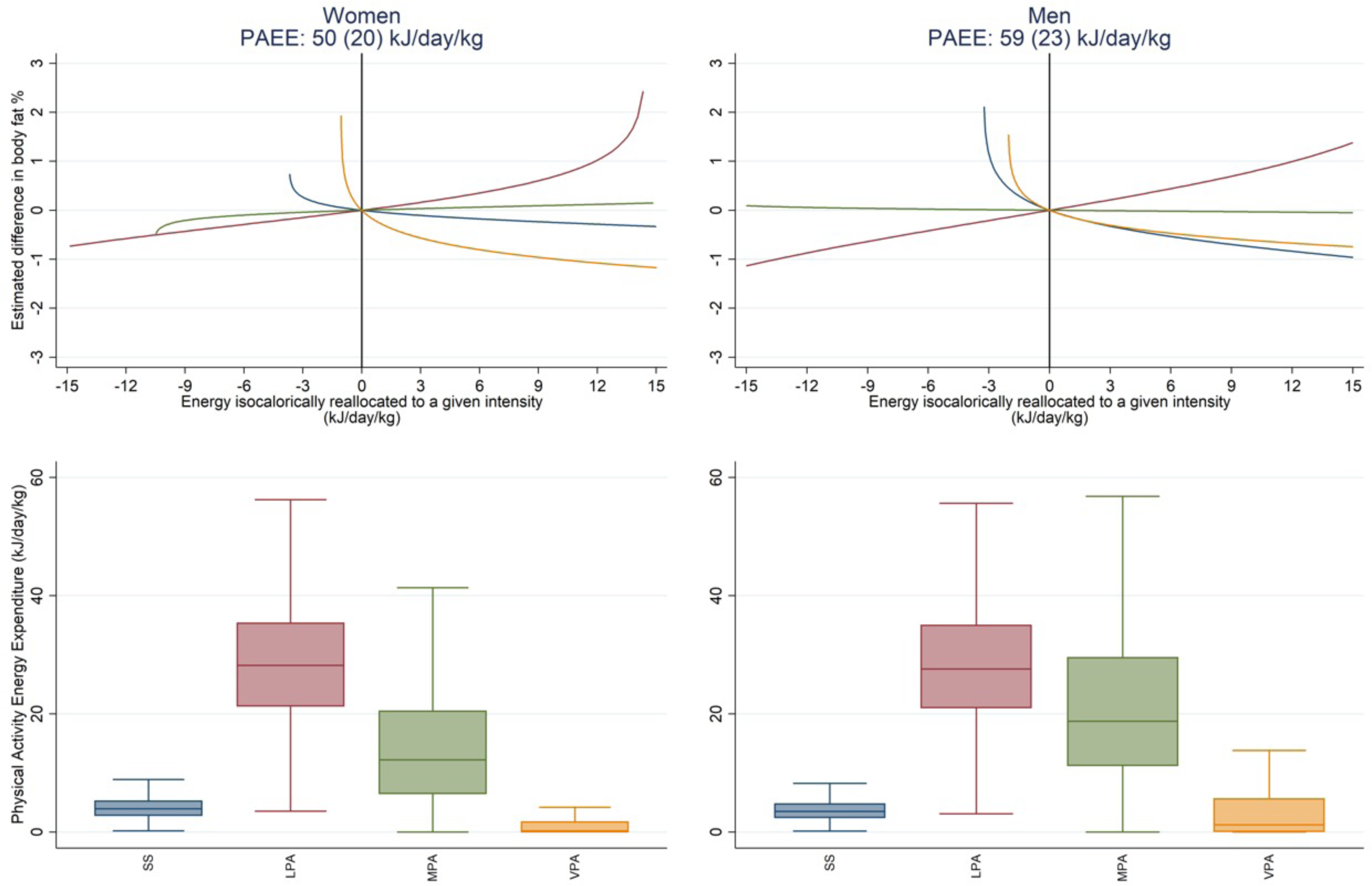
Estimated difference in body fat percentage associated with the isocaloric reallocation of PAEE to different intensities and box plots of the distribution of the PAEE composition, stratified by sex. The top panel shows the relative estimated difference in body fat percentage associated with an isocaloric reallocation of energy proportionately from all behaviours to the intensity of interest, as modelled by compositional data analysis. The origin (x=0,y=0) represents no change in the intensity composition of PAEE from the mean composition of the group of interest (women and men). The bottom panel illustrates the relative size of each reservoir of energy across women and men. Group PAEE values are mean (SD). SS = 0-1.5 METs, LPA = 1.5-3 METs, MPA = 3-6 METs, VPA >6 METs.

To model the specific reallocation of energy from one specific intensity to another, we also conducted pairwise compositional analysis. This analysis is presented in Figure S1, alongside a graphical depiction of the linear substitution model. The regression results underpinning these curves can be found in Supplementary Table 5.

To further explore the relative importance of intensity and volume, we conducted a sensitivity analysis of the compositional analysis, with intensity categories redefined as above. This analysis showed a slightly greater magnitude of association for MPA energy in women, but the association was non-significant in men as before. In contrast, the estimated difference in BF% per kJ·day^-1^·kg^-1^ active energy reallocated to VPA was greater (Figure S2, Supplementary Table 6).

The relative importance of volume and composition is highlighted in Figure 4 (coefficients in Supplementary Table 7), which stratified the relationship between compositional intensity and body fat percentage by tertile of PAEE. On average, body fat percentage in the bottom vs top tertile of PAEE was 41% vs 34% in women and 30% vs 25% in men. The estimated difference in body fatness from reallocating PAEE to VPA was preserved across tertiles but was comparatively much smaller than the estimated effect of the absolute PAEE level itself.

**Figure 4:**
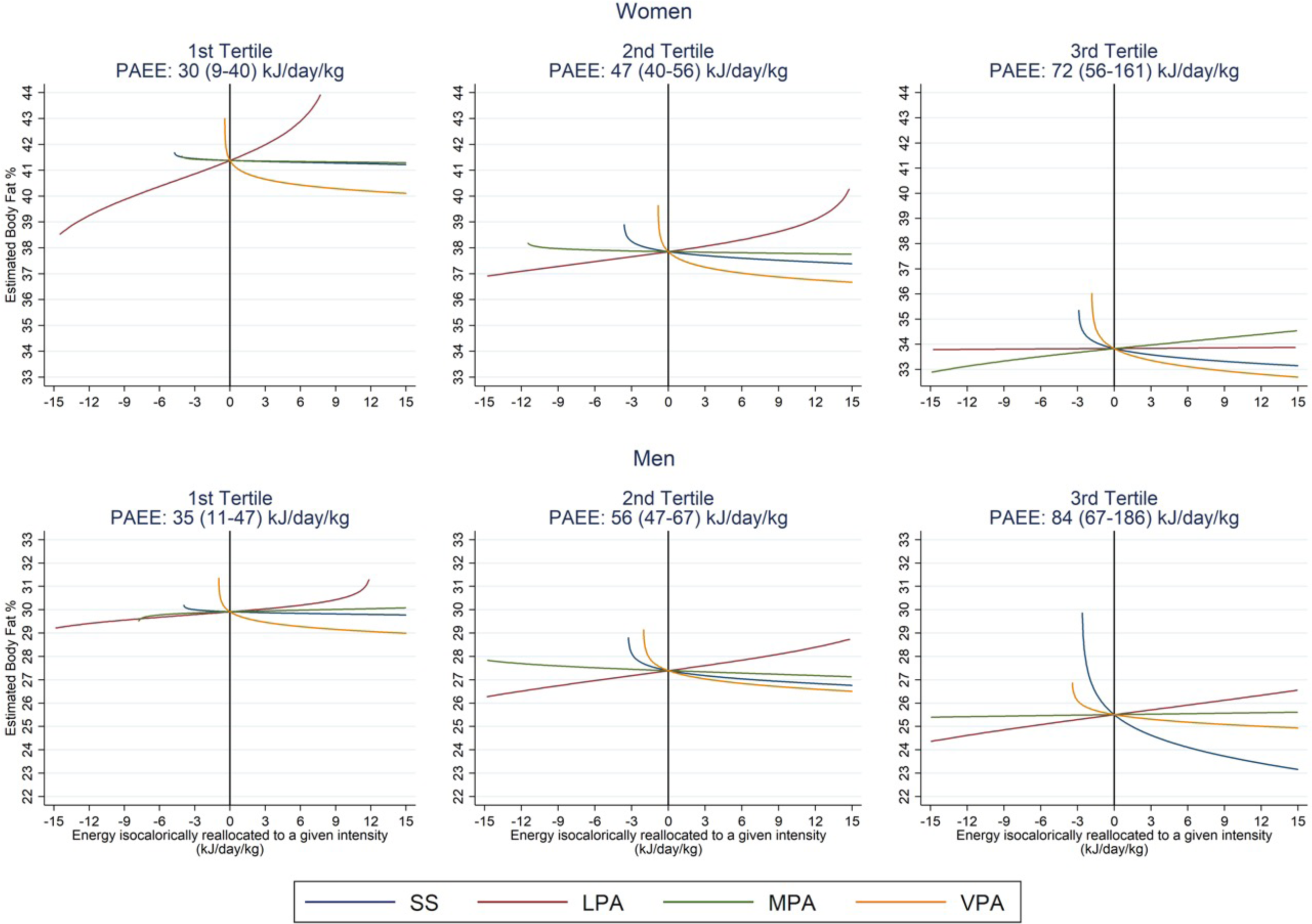
Estimated difference in body fat percentage associated with the isocaloric reallocation of PAEE to different intensities, stratified by sex and tertile of PAEE, as modelled by compositional data analysis. The origin (x=0,y=0) represents no change in the intensity composition of PAEE from the mean composition of the group of interest. Comparing groups by tertile at the intercept highlights the associated difference in body fat percentage attributable to varying levels of PAEE. Group PAEE values are mean (range). SS = 0-1.5 METs, LPA = 1.5-3 METs, MPA = 3-6 METs, VPA >6 METs.

For comparison, equivalent isotemporal associations between physical activity intensity and body composition are shown in Supplementary Figures S3 and S4. The regressions underpinning these graphs are found in Supplementary Tables 8, 9 and 10. These models estimate the effect of intensity reallocation in the time domain and are not controlled for overall PAEE volume.

## Discussion

In this large population-based cohort study of 11 468 free-living women and men with individually-calibrated, objectively-assessed physical activity and DXA-derived body composition, we show that overall physical activity volume is strongly inversely associated with body-fatness. Additionally, our integrated volume-and-intensity analyses demonstrate that higher proportions of more intense physical activity are associated with lower body-fatness, compared to when a similar volume of activity is accumulated at a lower intensity.

The association between overall PAEE and body-fatness is in keeping with other population studies that highlight the important role that PA plays in maintaining a healthy body weight^21–23^. Our study confirms this association across the middle years of adult life. Uniquely, we have also shown the relative importance of PAEE compared to intensity; this is particularly prominent in our analysis of intensity composition, stratified by PAEE tertile.

These results show that although more vigorous intensity activity is associated with lower body-fatness, the relative importance of intensity is smaller compared to the overarching contribution of PAEE volume. Indeed, all isocaloric associations must be considered in the context of overall PAEE. This is particularly important when considering the relative contribution of LPA, which was associated with higher body-fatness. In isolation, this result may appear paradoxical but when interpreted as a nested exposure within overall PAEE volume, it is not.

Given the contribution of LPA to overall PAEE, especially in women^11^, and the challenges that some people face in being active, for example due to advancing age, ailments or other factors^24^, the value of LPA should not be discounted. Indeed, assuming associations are causal, our results suggest that were a woman to increase her PAEE from the 1^st^ to the 2^nd^ tertile, an average increase of 17 kJ·day^-1^·kg^-1^, her BF% would be roughly 3 percentage points lower. This contrasts to approximately 1 percentage point lower BF% if her PAEE remained fixed, but she accumulated an additional 30% of that PAEE from VPA (the maximum percentage observed within this group). If she were to increase both volume and VPA as above, the combined estimated benefit would be 4.5 percentage points.

This is somewhat in contrast to the isotemporal substitution results, which may be perceived to have more direct relevance to the type of public health messaging which focuses on people’s time budget. These generally show that more time spent in higher intensity categories is associated with lower body fat. However, in the isotemporal analysis, the reallocation of time to higher intensity activity is associated with an inherent increase in volume. Thus, it is not possible to disentangle the estimated effects of volume and intensity. In addition, isotemporal analysis results also do not account for intensity variations within each intensity category, e.g. in our present analyses a 30-min activity at 3.1 METs is treated the same as a 30-min activity at 5.9 METs. In the integrated volume-and-intensity model in the energy space, this problem is effectively avoided as these two activities will result in substantively different PAEE levels as captured in the volume variable. This analysis acknowledges the nested nature of the exposures and aims to provide a simple answer to the primary question of whether overall physical activity is related to the outcome of interest, and secondarily what sub-dimensions of physical activity may or may not be associated beyond that.

Apart from the more biologically intuitive specification of such isocaloric models, the public health relevance of these models can be interpreted by recasting the issue of potential behaviour change away from time to considering life as a series of daily tasks. For example, a common daily task for most working people is their commute to work. Whilst desirable, it may not be possible for everyone to switch to greener and more active modes of transport for the entire journey, but it would be possible for most car drivers and people who use public transport to incorporate more walking by parking further away or getting off the bus one stop earlier. This would increase activity volume but would also increase pressure on the time budget unless also accompanied by an increase in intensity. Our results indicate clear benefits to body-fatness of both such behaviour changes. Were it not initially possible to increase activity volume this way, small upward adjustments to the intensity by which set tasks are undertaken, for example by walking a set distance a bit faster, would still yield some health benefits, and importantly, it would also create a time surplus. Over time this may be sufficient to also accommodate more significant volume changes for even greater health benefits. Conversely, any small changes in, for example, the built environment, which result in subtle intensity decreases of set physical tasks or indeed the abolition of some tasks altogether, may have negative consequences for population levels of obesity if the associations we report here are causal.

Our observations from this large cross-sectional study of UK adults are supported by evidence from trials seeking to delineate the contribution of activity intensity independent of PAEE. In a systematic review and meta-analysis of 28 intervention studies, Keating et al concluded that calorically matched high-intensity interval training and moderate-intensity continuous training provided similar benefits for reducing body fat, despite limitations of small sample sizes and incomplete control for confounding caused by variations in diet and physical activity outside of the training sessions that were part of the intervention^25^. A large population study such as ours provides evidence for the habitual physical activity perspective and has the advantage of being able to identify real-life activity profiles that associate with lower body-fatness, whilst controlling statistically for a range of confounders including habitual diet.

Our results, at least in the time domain, are also consistent with previous observations in older British adults employing a similar exposure estimation technique, in which a positive association for sedentary time and inverse associations for both LPA and MVPA on body-fatness were reported, with a stronger magnitude of association for MVPA^21^. Comparable associations were also reported in a small sample of older high-risk Spanish adults with metabolic syndrome using wrist accelerometry-derived physical activity and DXA-derived body-fatness as estimated using linear isotemporal substitution analysis^26^.

Likewise, an examination of BMI and waist circumference outcomes in US adults and the role of time spent in different physical activity intensity categories, modelled with compositional analysis, demonstrated an inverse cross-sectional association between time in MVPA and those outcomes^4^. Further, a recent longitudinal compositional study in elderly, Central European women, demonstrated that the longitudinal reallocation of time from MVPA to sedentary behaviour was associated with increased BMI and body-fatness^27^.

Our findings also line-up with recently reported associations between activity and all-cause mortality in the UK Biobank cohort^5^. These findings suggest that the health benefits of higher intensity of activity, while significant, were secondary to that of volume. Taken together, these findings support the recently released WHO guidelines, which, above all else, emphasise the notion that every move counts^28^.

Our study has several strengths including the large population size, the strength of the objective assessment methods for the exposures and outcomes, and the ability to control for a range of potential confounders. A further strength is the similarity of results across a range of modelling techniques. It is limited by its cross-sectional nature and further longitudinal studies and randomised controlled trials are needed before definitive statements about causality can be made.

## Conclusion

In this population-based study of objectively measured physical activity and body-fatness, our integrated analysis of activity volume and intensity show that total PAEE and the pattern of accumulation of PAEE were both significantly associated with body-fatness outcomes independent of one another. At similar levels of PAEE, a greater proportion of energy expended at a higher intensity is associated with lower body-fatness. However, this association is secondary in order of magnitude to that of overall volume.

## Data Availability

Contact the MRC Epidemiology Unit Cambridge with any data request.

## Authors’ contributions

The authors contributed to the present manuscript as follows: Idea for analysis (T.L., S.B.); acquisition, analysis of raw physical activity data (K.We., S.B.); acquisition, analysis of raw anthropometry and DXA data (E.D.L.R.) epidemiological data analysis (T.L.); drafting of the manuscript (T.L.); revising work critically for important intellectual content (all authors); approval of the final version before submission (all authors). Chief Investigator (N.J.W.) and Principal Investigators (N.G.F., S.G., S.B.) of the Fenland Study.

## Acknowledgements

We are grateful to the Fenland Study participants for their willingness and time to take part. The Fenland Study also has a dedicated Patient and Public Involvement panel, who provided input on the acceptability of the study protocols and the means by which participant data confidentiality is ensured. We thank all members of the following teams at the MRC Epidemiology Unit responsible for practical aspects of the study: Study Coordination, Field Epidemiology, Anthropometry Team, Physical Activity Technical Team, IT, Data Management, and Statistics. We would like to specifically acknowledge Stefanie Hollidge and Richard Powell for their assistance in the processing of physical activity and DXA data for this study.

## Supplement Legend

**Supplementary Table 1:**
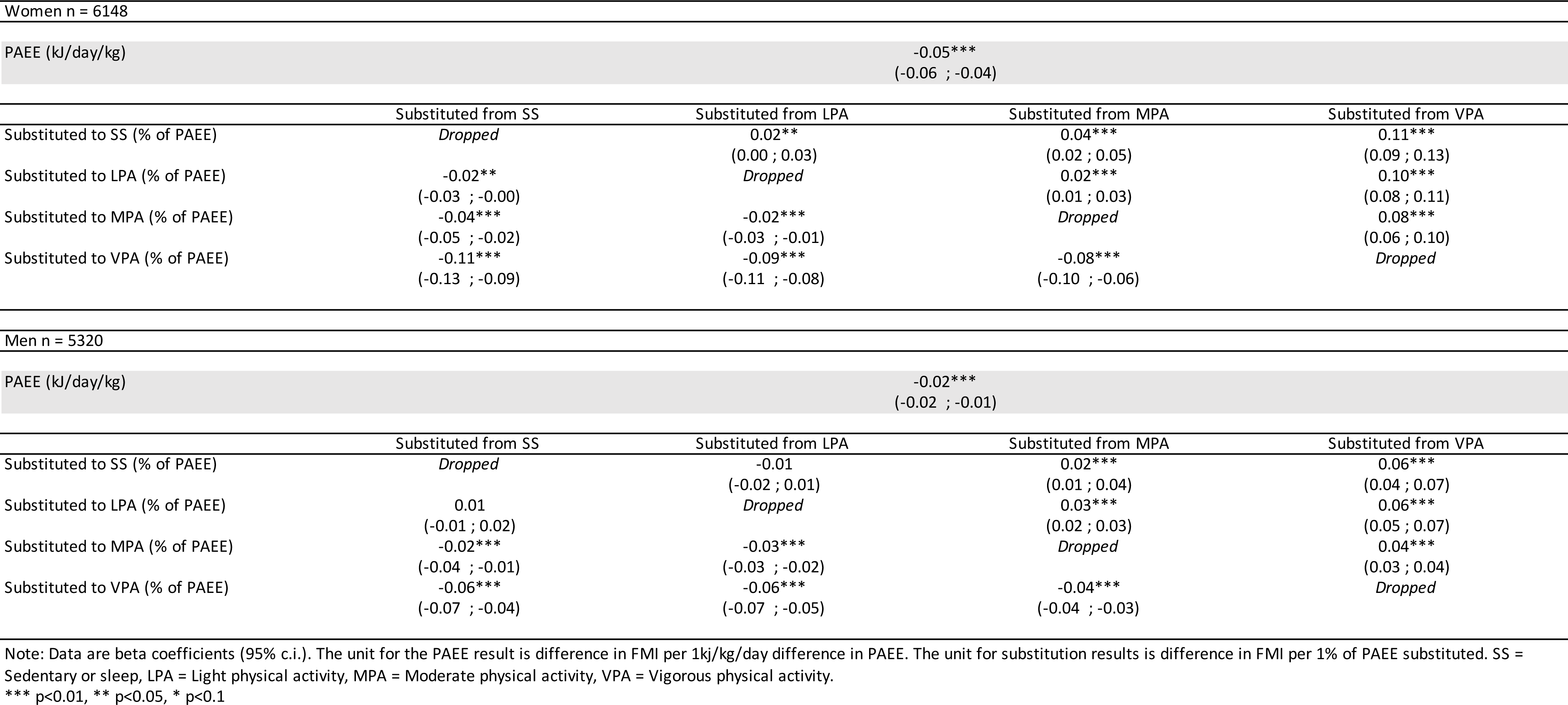
Isocaloric substitution of physical activity energy expenditure and fat mass index.

Women n = 6148
|  |  |  |  |  |
| --- | --- | --- | --- | --- |
| PAEE (kJ/day/kg) | -0.05***<br>(-0.06 ; -0.04) |  |  |  |
|  | Substituted from SS | Substituted from LPA | Substituted from MPA | Substituted from VPA |
| Substituted to SS (% of PAEE) | *Dropped* | 0.02**<br>(0.00 ; 0.03) | 0.04***<br>(0.02 ; 0.05) | 0.11***<br>(0.09 ; 0.13) |
| Substituted to LPA (% of PAEE) | -0.02**<br>(-0.03 ; -0.00) | *Dropped* | 0.02***<br>(0.01 ; 0.03) | 0.10***<br>(0.08 ; 0.11) |
| Substituted to MPA (% of PAEE) | -0.04***<br>(-0.05 ; -0.02) | -0.02***<br>(-0.03 ; -0.01) | *Dropped* | 0.08***<br>(0.06 ; 0.10) |
| Substituted to VPA (% of PAEE) | -0.11***<br>(-0.13 ; -0.09) | -0.09***<br>(-0.11 ; -0.08) | -0.08***<br>(-0.10 ; -0.06) | *Dropped* |
| Men n = 5320 |  |  |  |  |
| PAEE (kJ/day/kg) | -0.02***<br>(-0.02 ; -0.01) |  |  |  |
|  | Substituted from SS | Substituted from LPA | Substituted from MPA | Substituted from VPA |
| Substituted to SS (% of PAEE) | *Dropped* | -0.01<br>(-0.02 ; 0.01) | 0.02***<br>(0.01 ; 0.04) | 0.06***<br>(0.04 ; 0.07) |
| Substituted to LPA (% of PAEE) | 0.01<br>(-0.01 ; 0.02) | *Dropped* | 0.03***<br>(0.02 ; 0.03) | 0.06***<br>(0.05 ; 0.07) |
| Substituted to MPA (% of PAEE) | -0.02***<br>(-0.04 ; -0.01) | -0.03***<br>(-0.03 ; -0.02) | *Dropped* | 0.04***<br>(0.03 ; 0.04) |
| Substituted to VPA (% of PAEE) | -0.06***<br>(-0.07 ; -0.04) | -0.06***<br>(-0.07 ; -0.05) | -0.04***<br>(-0.04 ; -0.03) | *Dropped* |

Note: Data are beta coefficients (95% c.i.). The unit for the PAEE result is difference in FMI per 1kJ/kg/day difference in PAEE. The unit for substitution results is difference in FMI per 1% of PAEE substituted. SS = Sedentary or sleep, LPA = Light physical activity, MPA = Moderate physical activity, VPA = Vigorous physical activity.
\*\*\* p<0.01, \*\* p<0.05, \* p<0.1

**Supplementary Table 2:**
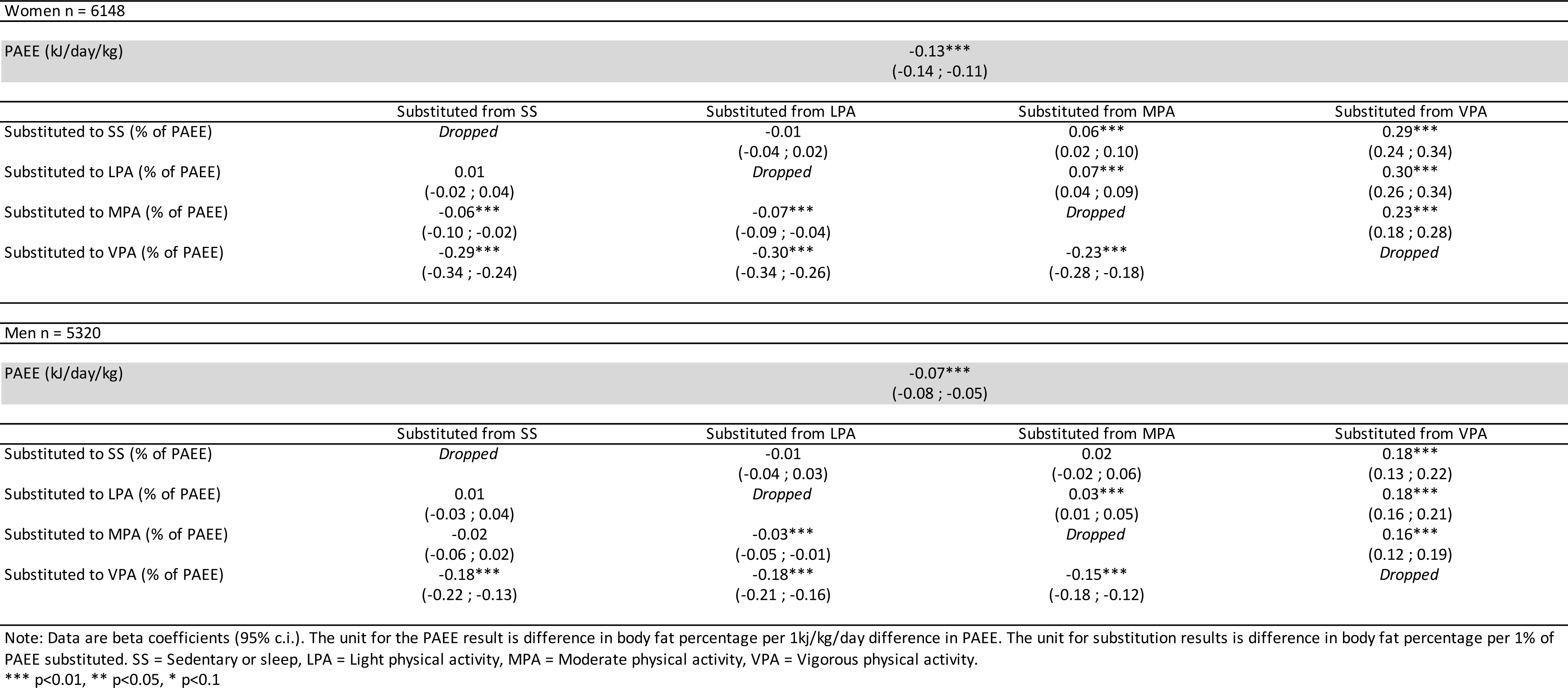
Isocaloric substitution of physical activity energy expenditure and body fat percentage using alternative intensity thresholds (MPA > 4 METs, VPA > 7METs).

**Supplementary Table 3:**
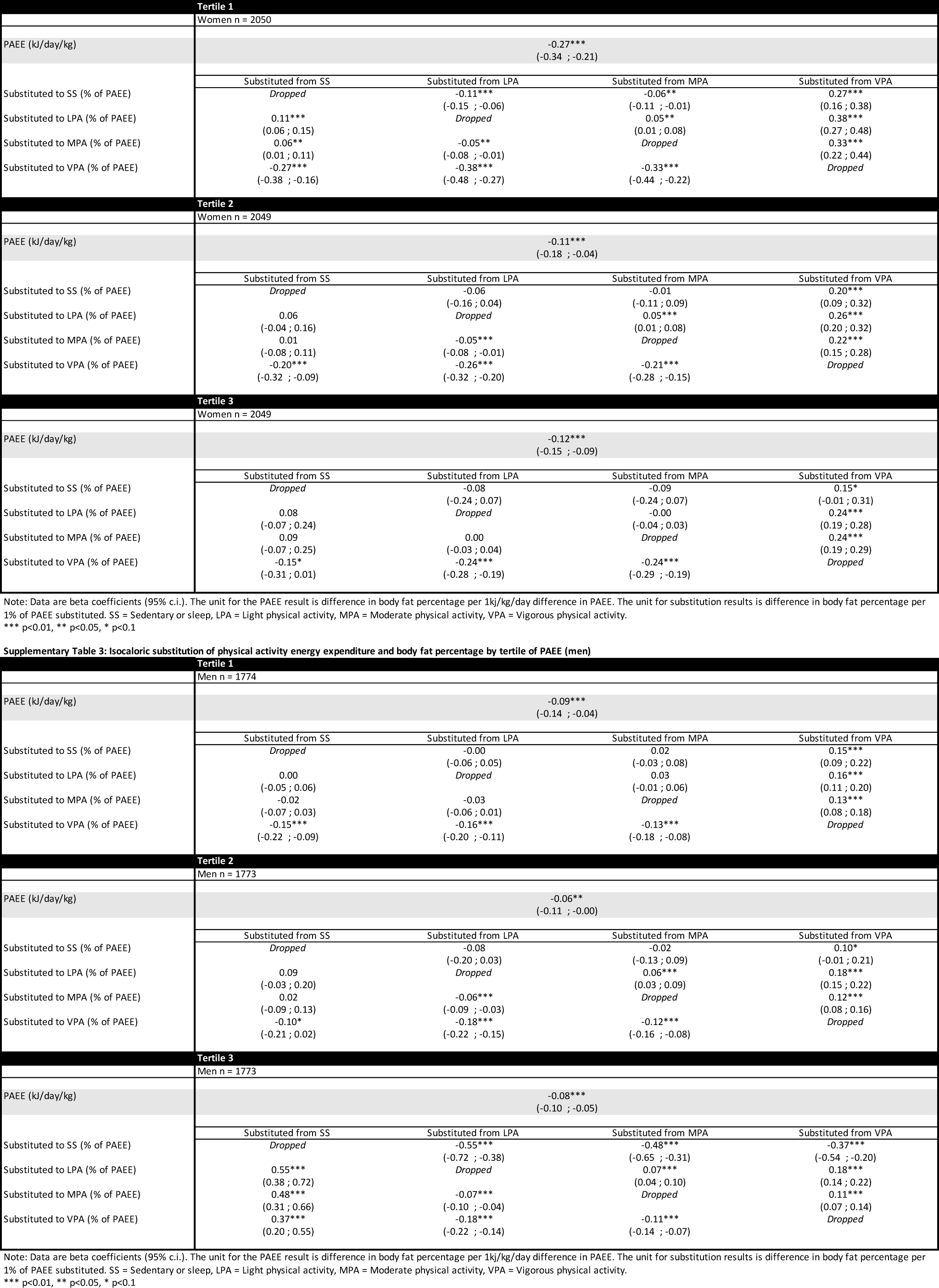
Isocaloric substitution of physical activity energy expenditure and body fat percentage by tertile of PAEE.

**Supplementary Table 4:** Relationship between isocaloric z1 ILR coordinate and body fat percentage.

| Women n = 6148 | coef | se | tstat | pval |
| --- | --- | --- | --- | --- |
| SS:remaining behaviours | -0.18* | (0.11) | -1.65 | 0.10 |
| LPA:remaining behaviours | 0.55*** | (0.14) | 4.02 | 0.00 |
| MPA:remaining behaviours | 0.11 | (0.10) | 1.07 | 0.29 |
| VPA:remaining behaviours | -0.48*** | (0.03) | -15.31 | 0.00 |
| Total PAEE | -0.13*** | (0.01) | -15.95 | 0.00 |
| Men n = 5320 | coef | se | tstat | pval |
| SS:remaining behaviours | -0.52*** | (0.10) | -5.15 | 0.00 |
| LPA:remaining behaviours | 0.95*** | (0.13) | 7.27 | 0.00 |
| MPA:remaining behaviours | -0.04 | (0.12) | -0.38 | 0.71 |
| VPA:remaining behaviours | -0.38*** | (0.03) | -13.23 | 0.00 |
| Total PAEE | -0.08*** | (0.01) | -11.51 | 0.00 |

Note: Beta coefficients speak to direction and significance, but not magnitude of reallocation effect. SS = Sedentary or sleep, LPA = Light physical activity, MPA = Moderate physical activity, VPA = Vigorous physical activity.
\*\*\* p<0.01, \*\* p<0.05, \* p<0.1

**Supplementary Table 5:** Relationship between isocaloric pairwise ILR coordinates and body fat percentage.

|  | Women n = 6148 |  |  |  | Men = 5320 |  |  |  |
| --- | --- | --- | --- | --- | --- | --- | --- | --- |
|  | coef | se | tstat | pval | coef | se | tstat | pval |
| **Physical Activity Energy Expenditure** | -0.13*** | (0.01) | -15.95 | 0.00 | -0.08*** | (0.01) | -11.51 | 0.00 |
| **Composition of intensity (Full ILR coordinate set)** | - | - | - | 0.00† | - | - | - | 0.00† |
| **Pairwise reallocation - SS to LPA** |  |  |  |  |  |  |  |  |
| $ilr_1 \propto \ln(\text{LPA} \& \text{SS}:\text{MPA} \& \text{VPA})$ | 0.56*** | (0.15) | 3.73 | 0.00 | 0.64*** | (0.17) | 3.81 | 0.00 |
| $ilr_2 \propto \ln(\text{LPA}:\text{SS})$ | 0.78*** | (0.24) | 3.23 | 0.00 | 1.56*** | (0.22) | 7.18 | 0.00 |
| $ilr_3 \propto \ln(\text{MPA}:\text{VPA})$ | 0.63*** | (0.12) | 5.07 | 0.00 | 0.36*** | (0.14) | 2.66 | 0.01 |
| **Pairwise reallocation - SS to MPA** |  |  |  |  |  |  |  |  |
| $ilr_1 \propto \ln(\text{MPA} \& \text{SS}:\text{LPA} \& \text{VPA})$ | -0.10 | (0.21) | -0.48 | 0.63 | -0.85*** | (0.20) | -4.17 | 0.00 |
| $ilr_2 \propto \ln(\text{MPA}:\text{SS})$ | 0.31* | (0.17) | 1.85 | 0.06 | 0.50*** | (0.18) | 2.78 | 0.01 |
| $ilr_3 \propto \ln(\text{LPA}:\text{VPA})$ | 1.10*** | (0.15) | 7.47 | 0.00 | 1.41*** | (0.14) | 10.10 | 0.00 |
| **Pairwise reallocation - SS to VPA** |  |  |  |  |  |  |  |  |
| $ilr_1 \propto \ln(\text{VPA} \& \text{SS}:\text{MPA} \& \text{LPA})$ | -0.99*** | (0.16) | -6.03 | 0.00 | -1.36*** | (0.16) | -8.72 | 0.00 |
| $ilr_2 \propto \ln(\text{VPA}:\text{SS})$ | -0.32*** | (0.12) | -2.60 | 0.01 | 0.14 | (0.11) | 1.28 | 0.20 |
| $ilr_3 \propto \ln(\text{MPA}:\text{LPA})$ | -0.47** | (0.23) | -2.03 | 0.04 | -1.05*** | (0.24) | -4.41 | 0.00 |
| **Pairwise reallocation - LPA to MPA** |  |  |  |  |  |  |  |  |
| $ilr_1 \propto \ln(\text{MPA} \& \text{LPA}:\text{SS} \& \text{VPA})$ | 0.99*** | (0.16) | 6.03 | 0.00 | 1.36*** | (0.16) | 8.72 | 0.00 |
| $ilr_2 \propto \ln(\text{MPA}:\text{LPA})$ | -0.47** | (0.23) | -2.03 | 0.04 | -1.05*** | (0.24) | -4.41 | 0.00 |
| $ilr_3 \propto \ln(\text{SS}:\text{VPA})$ | 0.32*** | (0.12) | 2.60 | 0.01 | -0.14 | (0.11) | -1.28 | 0.20 |
| **Pairwise reallocation - LPA to VPA** |  |  |  |  |  |  |  |  |
| $ilr_1 \propto \ln(\text{VPA} \& \text{LPA}:\text{SS} \& \text{MPA})$ | 0.10 | (0.21) | 0.48 | 0.63 | 0.85*** | (0.20) | 4.17 | 0.00 |
| $ilr_2 \propto \ln(\text{VPA}:\text{LPA})$ | -1.10*** | (0.15) | -7.47 | 0.00 | -1.41*** | (0.14) | -10.10 | 0.00 |
| $ilr_3 \propto \ln(\text{SS}:\text{MPA})$ | -0.31* | (0.17) | -1.85 | 0.06 | -0.50*** | (0.18) | -2.78 | 0.01 |
| **Pairwise reallocation - MPA to VPA** |  |  |  |  |  |  |  |  |
| $ilr_1 \propto \ln(\text{MPA} \& \text{VPA}:\text{SS} \& \text{LPA})$ | -0.56*** | (0.15) | -3.73 | 0.00 | -0.64*** | (0.17) | -3.81 | 0.00 |
| $ilr_2 \propto \ln(\text{VPA}:\text{MPA})$ | -0.63*** | (0.12) | -5.07 | 0.00 | -0.36*** | (0.14) | -2.66 | 0.01 |
| $ilr_3 \propto \ln(\text{SS}:\text{LPA})$ | -0.78*** | (0.24) | -3.23 | 0.00 | -1.56*** | (0.22) | -7.18 | 0.00 |

\*\*\* p<0.01, \*\* p<0.05, \* p<0.1. SS = Sedentary or sleep, LPA = Light physical activity, MPA = Moderate physical activity, VPA = Vigorous physical activity.
† Likelihood ratio test for contribution of relative intensity composition

**Supplementary Table 6:** Relationship between isocaloric z1 ILR coordinate and body fat percentage (MPA > 4MET, VPA > 7METs).

| Women n = 6148 | coef | se | tstat | pval |
| --- | --- | --- | --- | --- |
| SS:remaining behaviours | -0.18* | (0.11) | -1.65 | 0.10 |
| LPA:remaining behaviours | 0.89*** | (0.12) | 7.46 | 0.00 |
| MPA:remaining behaviours | -0.23*** | (0.05) | -4.33 | 0.00 |
| VPA:remaining behaviours | -0.48*** | (0.03) | -14.44 | 0.00 |
| Total PAEE | -0.12*** | (0.01) | -15.44 | 0.00 |

| Men n = 5320 | coef | se | tstat | pval |
| --- | --- | --- | --- | --- |
| SS:remaining behaviours | -0.48*** | (0.10) | -4.64 | 0.00 |
| LPA:remaining behaviours | 0.91*** | (0.12) | 7.86 | 0.00 |
| MPA:remaining behaviours | -0.04 | (0.07) | -0.65 | 0.52 |
| VPA:remaining behaviours | -0.39*** | (0.03) | -13.53 | 0.00 |
| Total PAEE | -0.08*** | (0.01) | -12.94 | 0.00 |

Note: Beta coefficients speak to direction and significance, but not magnitude of reallocation effect. SS = Sedentary or sleep, LPA = Light physical activity, MPA = Moderate physical activity, VPA = Vigorous physical activity.
\*\*\* p<0.01, \*\* p<0.05, \* p<0.1

**Supplementary Table 7:**
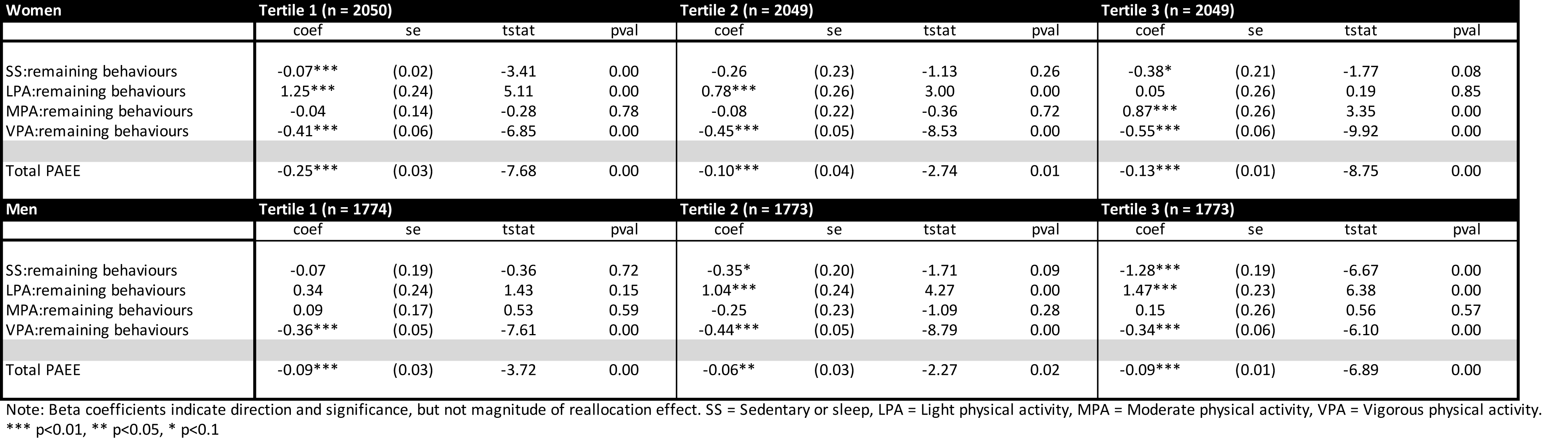
Relationship between isocaloric z1 ILR coordinate and body fat percentage by tertile of PAEE.

**Supplementary Table 8:** Isotemporal substitution of physical activity and body fat percentage.

Women n = 6148
|  | Substituted from SS | Substituted from LPA | Substituted from MPA | Substituted from VPA |
| --- | --- | --- | --- | --- |
| Substituted to SS (min/day) | *Dropped* | 0.01***<br>(0.01 - 0.01) | 0.03***<br>(0.03 - 0.03) | 0.22***<br>(0.20 - 0.25) |
| Substituted to LPA (min/day) | -0.01***<br>(-0.01 - -0.01) | *Dropped* | 0.02***<br>(0.02 - 0.02) | 0.21***<br>(0.19 - 0.24) |
| Substituted to MPA (min/day) | -0.03***<br>(-0.03 - -0.03) | -0.02***<br>(-0.02 - -0.02) | *Dropped* | 0.19***<br>(0.17 - 0.22) |
| Substituted to VPA (min/day) | -0.22***<br>(-0.25 - -0.20) | -0.21***<br>(-0.24 - -0.19) | -0.19***<br>(-0.22 - -0.17) | *Dropped* |

Men n = 5320
|  | Substituted from SS | Substituted from LPA | Substituted from MPA | Substituted from VPA |
| --- | --- | --- | --- | --- |
| Substituted to SS (min/day) | *Dropped* | -0.00***<br>(-0.00 - -0.00) | 0.02***<br>(0.01 - 0.02) | 0.12***<br>(0.10 - 0.13) |
| Substituted to LPA (min/day) | 0.00***<br>(0.00 - 0.00) | *Dropped* | 0.02***<br>(0.02 - 0.02) | 0.12***<br>(0.11 - 0.13) |
| Substituted to MPA (min/day) | -0.02***<br>(-0.02 - -0.01) | -0.02***<br>(-0.02 - -0.02) | *Dropped* | 0.10***<br>(0.09 - 0.12) |
| Substituted to VPA (min/day) | -0.12***<br>(-0.13 - -0.10) | -0.12***<br>(-0.13 - -0.11) | -0.10***<br>(-0.12 - -0.09) | *Dropped* |

Note: Data are beta coefficients (95% c.i.). The unit for substitution results is difference in body fat percentage per minute per day substituted. SS = Sedentary or sleep, LPA = Light physical activity, MPA = Moderate physical activity, VPA = Vigorous physical activity.
\*\*\* p<0.01, \*\* p<0.05, \* p<0.1

**Supplementary Table 9:** Relationship between isotemporal z1 ILR coordinate and body fat percentage.

| Women n = 6148 | coef | se | tstat | pval |
| --- | --- | --- | --- | --- |
| SS:remaining behaviours | 1.81*** | (0.14) | 12.94 | 0.00 |
| LPA:remaining behaviours | -0.45*** | (0.17) | -2.66 | 0.01 |
| MPA:remaining behaviours | -0.90*** | (0.08) | -10.99 | 0.00 |
| VPA:remaining behaviours | -0.46*** | (0.02) | -18.59 | 0.00 |

| Men n = 5320 | coef | se | tstat | pval |
| --- | --- | --- | --- | --- |
| SS:remaining behaviours | 0.40*** | (0.14) | 2.94 | 0.00 |
| LPA:remaining behaviours | 0.93*** | (0.15) | 6.12 | 0.00 |
| MPA:remaining behaviours | -0.99*** | (0.09) | -11.28 | 0.00 |
| VPA:remaining behaviours | -0.34*** | (0.02) | -14.34 | 0.00 |

Note: Beta coefficients speak to direction and significance, but not magnitude of reallocation effect. SS = Sedentary or sleep, LPA = Light physical activity, MPA = Moderate physical activity, VPA = Vigorous physical activity.
\*\*\* p<0.01, \*\* p<0.05, \* p<0.1

**Supplementary Table 10:** Relationship between isotemporal pairwise ILR coordinates and body fat percentage.

|  | Women n = 6148 |  |  |  | Men = 5320 |  |  |  |
| --- | --- | --- | --- | --- | --- | --- | --- | --- |
|  | coef | se | tstat | pval | coef | se | tstat | pval |
| **Composition of intensity (Full ILR coordinate set)** | - | - | - | 0.00† | - | - | - | 0.00† |
| **Pairwise reallocation - SS to LPA** |  |  |  |  |  |  |  |  |
| $ilr_1 \propto \ln(\text{LPA} \& \text{SS}:\text{MPA} \& \text{VPA})$ | 2.04*** | (0.11) | 18.24 | 0.00 | 2.00*** | (0.12) | 16.43 | 0.00 |
| $ilr_2 \propto \ln(\text{LPA}:\text{SS})$ | -2.39*** | (0.32) | -7.51 | 0.00 | 0.56* | (0.29) | 1.92 | 0.05 |
| $ilr_3 \propto \ln(\text{MPA}:\text{VPA})$ | -0.47*** | (0.10) | -4.67 | 0.00 | -0.68*** | (0.11) | -6.47 | 0.00 |
| **Pairwise reallocation - SS to MPA** |  |  |  |  |  |  |  |  |
| $ilr_1 \propto \ln(\text{MPA} \& \text{SS}:\text{LPA} \& \text{VPA})$ | 1.35*** | (0.26) | 5.25 | 0.00 | -0.88*** | (0.23) | -3.75 | 0.00 |
| $ilr_2 \propto \ln(\text{MPA}:\text{SS})$ | -2.87*** | (0.16) | -17.91 | 0.00 | -1.47*** | (0.18) | -8.32 | 0.00 |
| $ilr_3 \propto \ln(\text{LPA}:\text{VPA})$ | 0.01 | (0.18) | 0.05 | 0.96 | 1.35*** | (0.16) | 8.43 | 0.00 |
| **Pairwise reallocation - SS to VPA** |  |  |  |  |  |  |  |  |
| $ilr_1 \propto \ln(\text{VPA} \& \text{SS}:\text{MPA} \& \text{LPA})$ | 2.03*** | (0.21) | 9.56 | 0.00 | 0.08 | (0.21) | 0.41 | 0.68 |
| $ilr_2 \propto \ln(\text{VPA}:\text{SS})$ | -2.40*** | (0.15) | -15.91 | 0.00 | -0.79*** | (0.15) | -5.34 | 0.00 |
| $ilr_3 \propto \ln(\text{MPA}:\text{LPA})$ | -0.48** | (0.24) | -2.03 | 0.04 | -2.04*** | (0.22) | -9.27 | 0.00 |
| **Pairwise reallocation - LPA to MPA** |  |  |  |  |  |  |  |  |
| $ilr_1 \propto \ln(\text{MPA} \& \text{LPA}:\text{SS} \& \text{VPA})$ | -2.03*** | (0.21) | -9.56 | 0.00 | -0.08 | (0.21) | -0.41 | 0.68 |
| $ilr_2 \propto \ln(\text{MPA}:\text{LPA})$ | -0.48** | (0.24) | -2.03 | 0.04 | -2.04*** | (0.22) | -9.27 | 0.00 |
| $ilr_3 \propto \ln(\text{SS}:\text{VPA})$ | 2.40*** | (0.15) | 15.91 | 0.00 | 0.79*** | (0.15) | 5.34 | 0.00 |
| **Pairwise reallocation - LPA to VPA** |  |  |  |  |  |  |  |  |
| $ilr_1 \propto \ln(\text{VPA} \& \text{LPA}:\text{SS} \& \text{MPA})$ | -1.35*** | (0.26) | -5.25 | 0.00 | 0.88*** | (0.23) | 3.75 | 0.00 |
| $ilr_2 \propto \ln(\text{VPA}:\text{LPA})$ | -0.01 | (0.18) | -0.05 | 0.96 | -1.35*** | (0.16) | -8.43 | 0.00 |
| $ilr_3 \propto \ln(\text{SS}:\text{MPA})$ | 2.87*** | (0.16) | 17.91 | 0.00 | 1.47*** | (0.18) | 8.32 | 0.00 |
| **Pairwise reallocation - MPA to VPA** |  |  |  |  |  |  |  |  |
| $ilr_1 \propto \ln(\text{MPA} \& \text{VPA}:\text{SS} \& \text{LPA})$ | -2.04*** | (0.11) | -18.24 | 0.00 | -2.00*** | (0.12) | -16.43 | 0.00 |
| $ilr_2 \propto \ln(\text{VPA}:\text{MPA})$ | 0.47*** | (0.10) | 4.67 | 0.00 | 0.68*** | (0.11) | 6.47 | 0.00 |
| $ilr_3 \propto \ln(\text{SS}:\text{LPA})$ | 2.39*** | (0.32) | 7.51 | 0.00 | -0.56* | (0.29) | -1.92 | 0.05 |

\*\*\* p<0.01, \*\* p<0.05, \* p<0.1. SS = Sedentary or sleep, LPA = Light physical activity, MPA = Moderate physical activity, VPA = Vigorous physical activity.
† Likelihood ratio test for contribution of relative intensity composition

**Figure S1:**
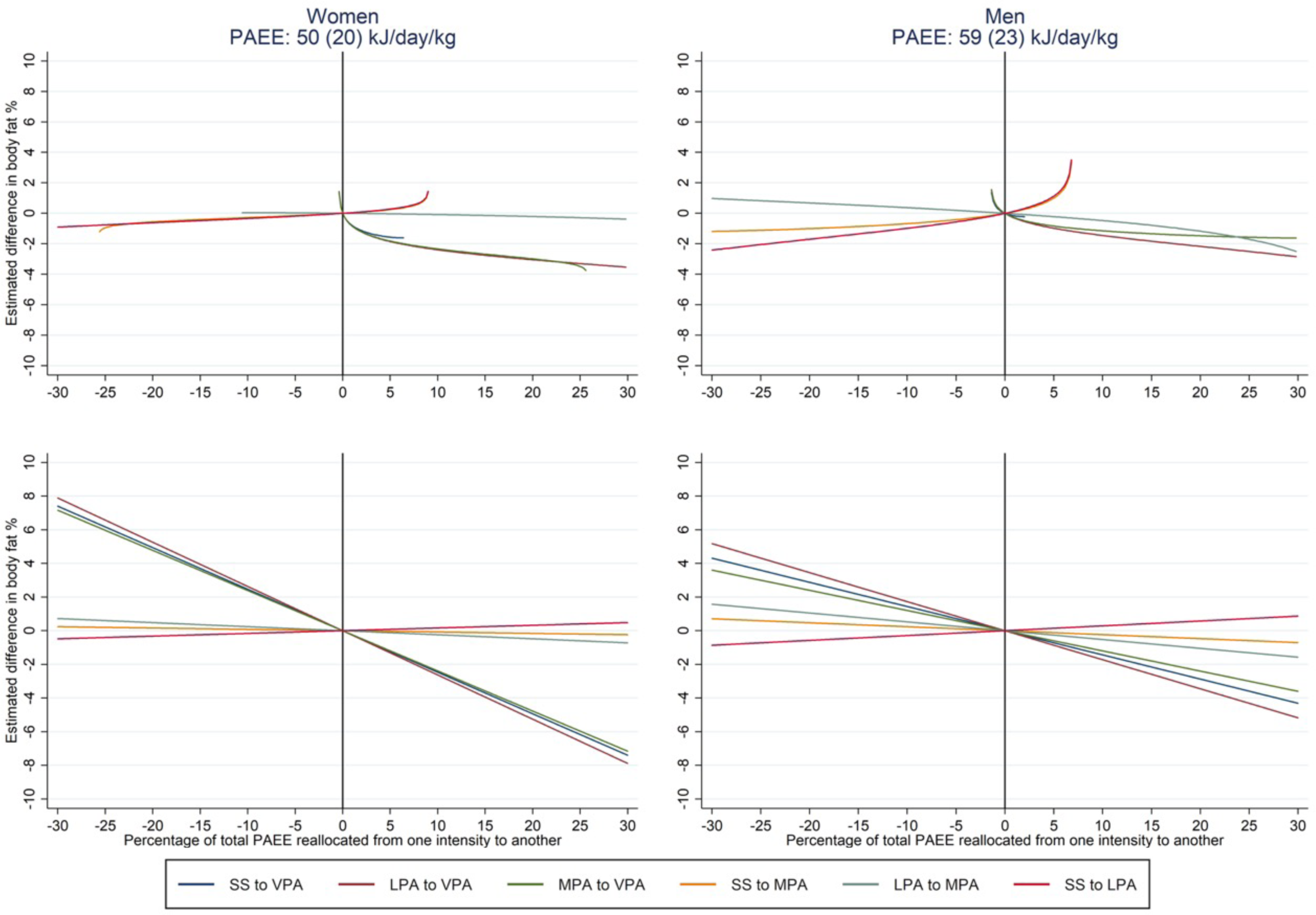
Pairwise and linear isocaloric reallocation of PAEE from one intensity to another. The top panel shows the results of pairwise compositional analysis, whereas the bottom panel shows linear substitution analysis. Both models estimate the difference in body fat percentage per 1% of PAEE reallocated. Group PAEE values are mean (SD). SS = 0-1.5 METs, LPA = 1.5-3 METs, MPA = 3-6 METs, VPA >6 METs.

**Figure S2:**
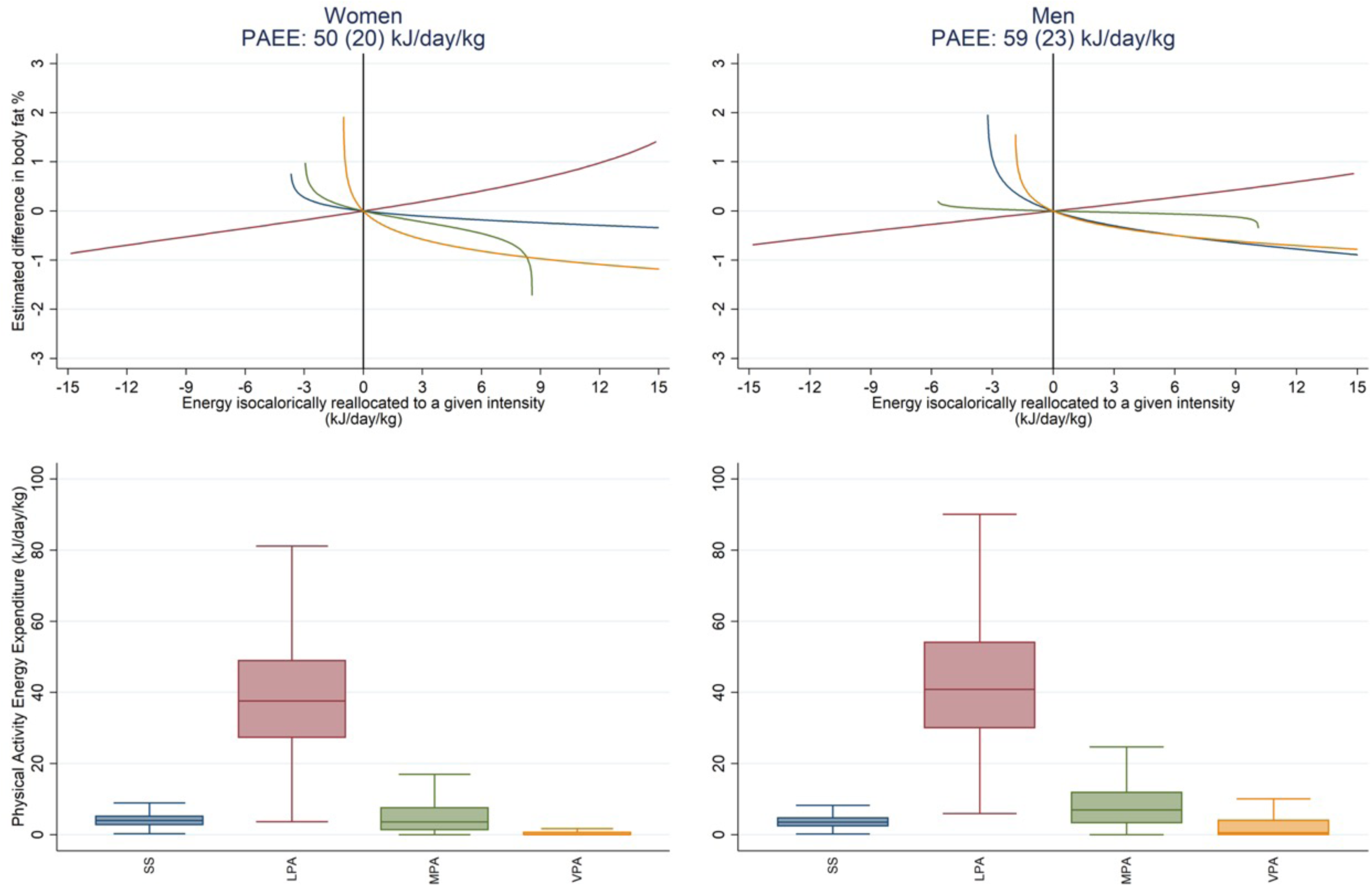
Estimated difference in body fat % associated with the isocaloric reallocation of PAEE to different intensities and box plots of the distribution of the PAEE composition, stratified by sex. Intensity thresholds redefined as SS = 0-1.5 METs, LPA = 1.5-4 METs, MPA = 4-7 METs, VPA >7 METs. The top panel shows the relative estimated difference in body fat percentage associated with an isocaloric reallocation of energy proportionately from all behaviours to the intensity of interest, as modelled by compositional data analysis. The origin (x=0,y=0) represents no change in the intensity composition of PAEE from the mean composition of the group of interest (women and men). The bottom panel illustrates the relative size of each reservoir of energy across women and men. Group PAEE values are mean (SD).

**Figure S3:**
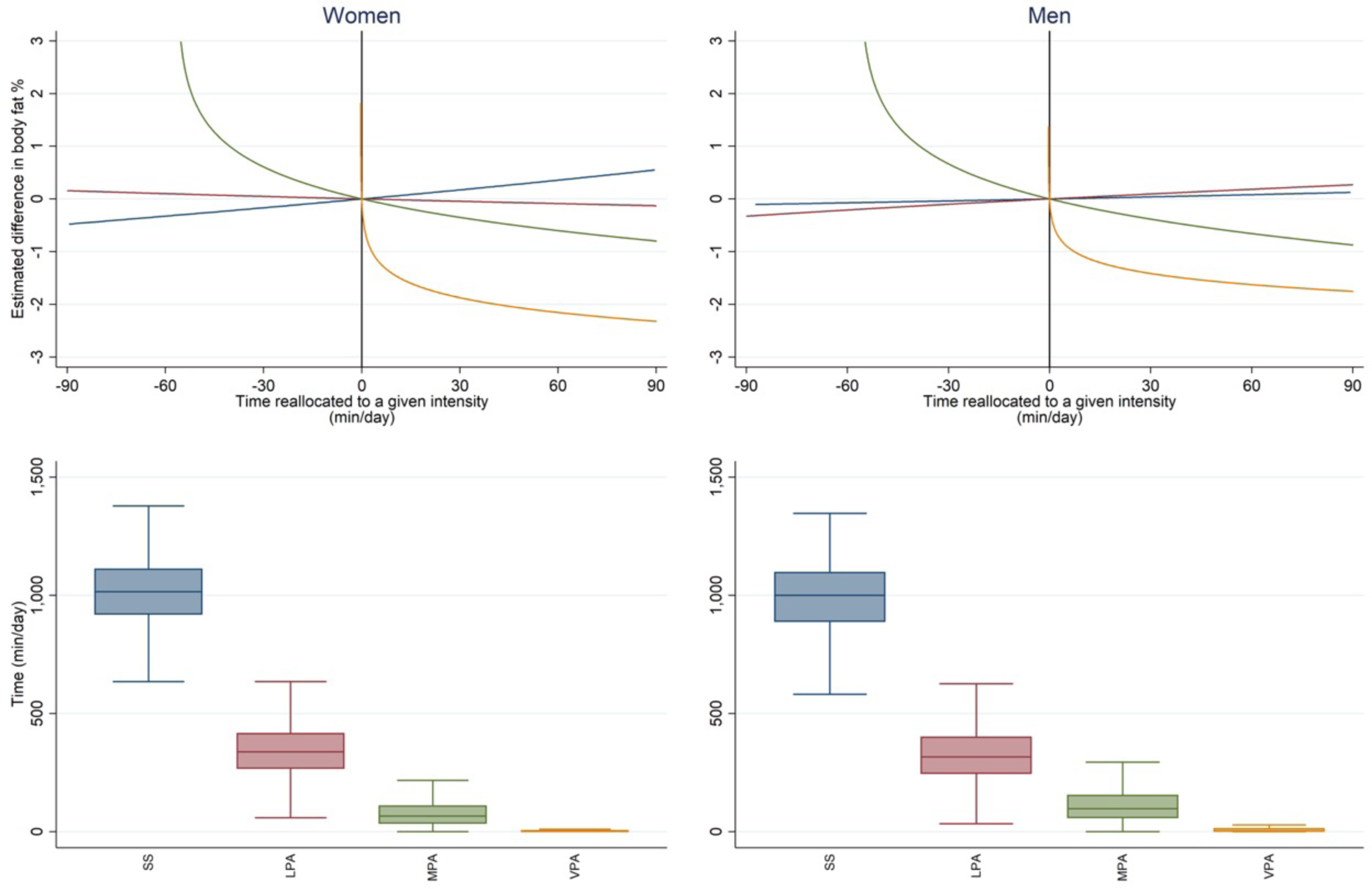
Estimated difference in body fat percentage associated with the isotemporal reallocation of time to different intensities, and box plots of the distribution of time by intensities, stratified by sex. The top panel shows the relative estimated difference in body fat percentage associated with the reallocation of time proportionately from all behaviours to the intensity of interest, as modelled by compositional data analysis. The origin (x=0,y=0) represents no change in the intensity composition of time from the mean composition of the group of interest (women and men). The bottom panel illustrates the relative size of each reservoir of time across women and men. SS = 0-1.5 METs, LPA = 1.5-3 METs, MPA = 3-6 METs, VPA >6 METs.

**Figure S4:**
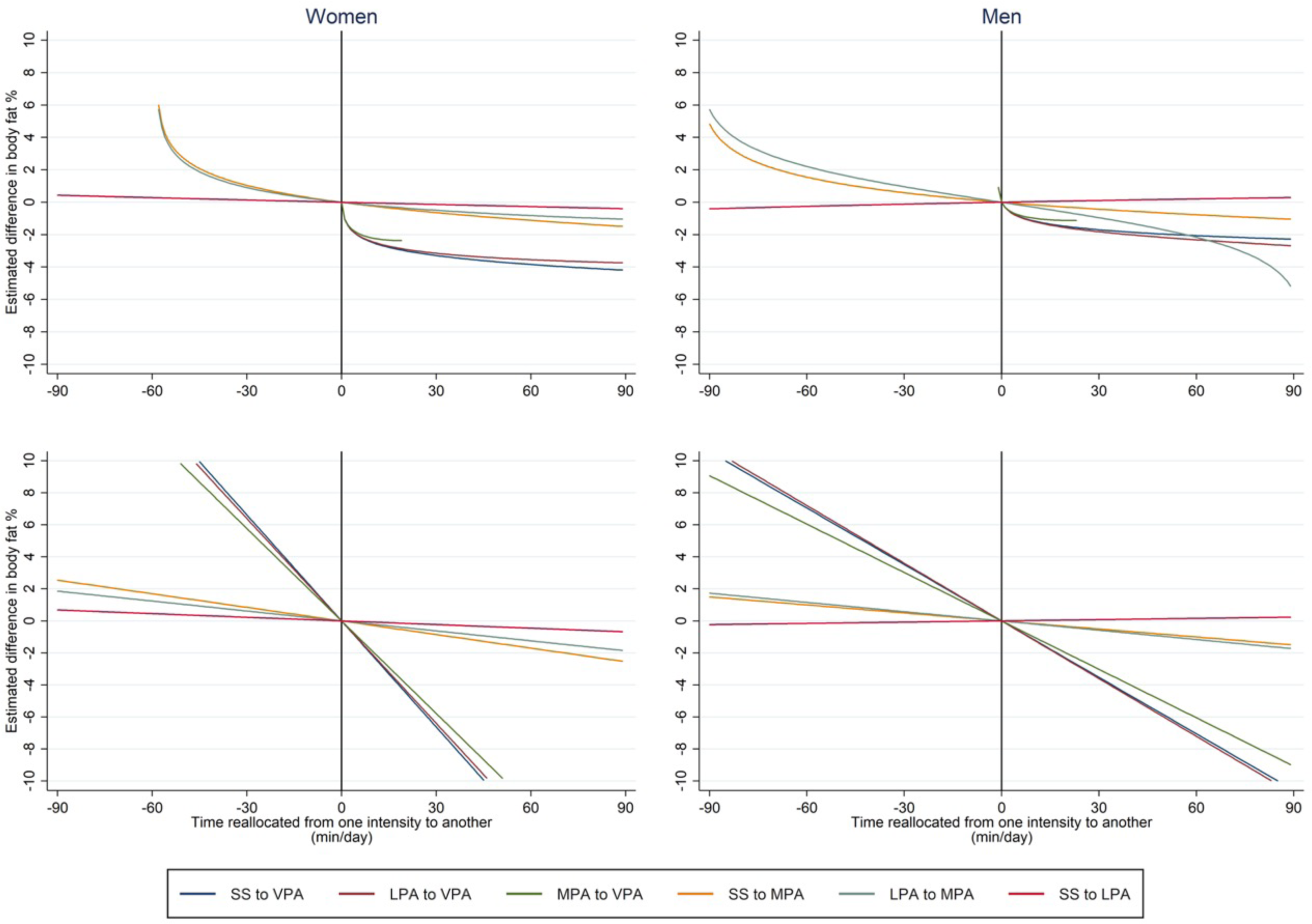
Pairwise and linear isotemporal reallocation of time from one intensity to another. The top panel shows the results of pairwise compositional analysis, whereas the bottom panel shows linear substitution analysis. Both models estimate the percentage difference in body fat % per minute per day reallocated. SS = 0-1.5 METs, LPA = 1.5-3 METs, MPA = 3-6 METs, VPA >6 METs.

## References

1. Smith AD, Crippa A, Woodcock J, Brage S. Physical activity and incident type 2 diabetes mellitus: a systematic review and dose–response meta-analysis of prospective cohort studies. Diabetologia 2016; 59: 2527–2545.

2. Moore SC, Lee I-M, Weiderpass E, Campbell PT, Sampson JN, Kitahara CM et al. Association of Leisure-Time Physical Activity With Risk of 26 Types of Cancer in 1.44 Million Adults. Jama Intern Med 2016; 176: 816.

3. Pearce M, Strain T, Kim Y, Sharp SJ, Westgate K, Wijndaele K, et al. Estimating physical activity from self-reported behaviours in large-scale population studies using network harmonisation: findings from UK Biobank and associations with disease outcomes. Int J Behav Nutr Phy 2020; 17: 40.

4. Chastin SFM, Palarea-Albaladejo J, Dontje ML, Skelton DA. Combined Effects of Time Spent in Physical Activity, Sedentary Behaviors and Sleep on Obesity and Cardio-Metabolic Health Markers: A Novel Compositional Data Analysis Approach. Plos One 2015; 10: e0139984.

5. Strain T, Wijndaele K, Dempsey PC, Sharp SJ, Pearce M, Jeon J et al. Wearable-device-measured physical activity and future health risk. Nat Med 2020;: 1–7.

6. Dumuid D, Pedišić Ž, Stanford TE, Martín-Fernández J-A, Hron K, Maher CA et al. The compositional isotemporal substitution model: A method for estimating changes in a health outcome for reallocation of time between sleep, physical activity and sedentary behaviour. Stat Methods Med Res 2017; 28: 846–857.

7. Dumuid D, Stanford TE, Martin-Fernández J-A, Pedišić Ž, Maher CA, Lewis LK et al. Compositional data analysis for physical activity, sedentary time and sleep research. Stat Methods Med Res 2017; 27: 3726–3738.

8. Wijndaele K, White T, Andersen LB, Bugge A, Kolle E, Northstone K, et al. Substituting prolonged sedentary time and cardiovascular risk in children and youth: a meta-analysis within the International Children’s Accelerometry database (ICAD). Int J Behav Nutr Phy 2019; 16: 96.

9. Li SX, Imamura F, Schulze MB, Zheng J, Ye Z, Agudo A et al. Interplay between genetic predisposition, macronutrient intake and type 2 diabetes incidence: analysis within EPIC-InterAct across eight European countries. Diabetologia 2018; 61: 1325–1332.

10. Willett WC, Howe GR, Kushi LH. Adjustment for total energy intake in epidemiologic studies. Am J Clin Nutrition 1997; 65: 1220S–1228S.

11. Lindsay T, Westgate K, Wijndaele K, Hollidge S, Kerrison N, Forouhi N, et al. Descriptive epidemiology of physical activity energy expenditure in UK adults (The Fenland study). Int J Behav Nutr Phy 2019; 16: 126.

12. Watson LPE, Venables MC, Murgatroyd PR. An Investigation Into the Differences in Bone Density and Body Composition Measurements Between 2 GE Lunar Densitometers and Their Comparison to a 4-Component Model. J Clin Densitom 2017; 20: 498–506.

13. Brage S, Brage N, Franks PW, Ekelund U, Wareham NJ. Reliability and validity of the combined heart rate and movement sensor Actiheart. Eur J Clin Nutr 2005; 59: 1602118.

14. Brage S, Ekelund U, Brage N, Hennings MA, Froberg K, Franks PW et al. Hierarchy of individual calibration levels for heart rate and accelerometry to measure physical activity. J Appl Physiol 2007; 103: 682–692.

15. Stegle O, Fallert SV, MacKay DJ, Brage S. Gaussian Process Robust Regression for Noisy Heart Rate Data. Ieee T Bio-med Eng 2008; 55: 2143–2151.

16. Brage S, Brage N, Franks PW, Ekelund U, Wong M-Y, Andersen LB et al. Branched equation modeling of simultaneous accelerometry and heart rate monitoring improves estimate of directly measured physical activity energy expenditure. J Appl Physiol 2004; 96: 343–351.

17. Brage S, Westgate K, Franks PW, Stegle O, Wright A, Ekelund U et al. Estimation of Free-Living Energy Expenditure by Heart Rate and Movement Sensing: A Doubly-Labelled Water Study. Plos One 2015; 10: e0137206.

18. Bingham SA, Welch AA, McTaggart A, Mulligan AA, Runswick SA, Luben R et al. Nutritional methods in the European Prospective Investigation of Cancer in Norfolk. Public Health Nutr 2001; 4: 847–858.

19. Bingham S. Validation of dietary assessment methods in the UK arm of EPIC using weighed records, and 24-hour urinary nitrogen and potassium and serum vitamin C and carotenoids as biomarkers. Int J Epidemiol 1997; 26: 137S – 151.

20. Mekary RA, Willett WC, Hu FB, Ding EL. Isotemporal Substitution Paradigm for Physical Activity Epidemiology and Weight Change. Am J Epidemiol 2009; 170: 519–527.

21. Bann D, Kuh D, Wills AK, Adams J, Brage S, Cooper R et al. Physical Activity Across Adulthood in Relation to Fat and Lean Body Mass in Early Old Age: Findings From the Medical Research Council National Survey of Health and Development, 1946–2010. Am J Epidemiol 2014; 179: 1197–1207.

22. Brage S, Lindsay T, Venables M, Wijndaele K, Westgate K, Collins D et al. Descriptive epidemiology of energy expenditure in the UK: findings from the National Diet and Nutrition Survey 2008-15. Int J Epidemiol 2020; 49: 1007–1021.

23. Ekelund U, Brage S, Franks PW, Hennings S, Emms S, Wong M-Y et al. Physical activity energy expenditure predicts changes in body composition in middle-aged healthy whites: effect modification by age. Am J Clin Nutrition 2005; 81: 964–969.

24. Bredland EL, Söderström S, Vik K. Challenges and motivators to physical activity faced by retired men when ageing: a qualitative study. Bmc Public Health 2018; 18: 627.

25. Keating SE, Johnson NA, Mielke GI, Coombes JS. A systematic review and meta-analysis of interval training versus moderate-intensity continuous training on body adiposity. Obes Rev 2017; 18: 943–964.

26. Galmes-Panades AM, Konieczna J, Varela-Mato V, Abete I, Babio N, Fiol M et al. Changes in physical activity, sedentary behaviour and body composition: longitudinal analysis in the PREDIMED)-Plus trial. undefined 2020. doi:rs.3.rs-44115/v1.

27. Pelclová J, Štefelová N, Dumuid D, Pedišić Ž, Hron K, Gába A et al. Are longitudinal reallocations of time between movement behaviours associated with adiposity among elderly women? A compositional isotemporal substitution analysis. Int J Obesity 2020; 44: 1–8.

28. Bull FC, Al-Ansari SS, Biddle S, Borodulin K, Buman MP, Cardon G et al. World Health Organization 2020 guidelines on physical activity and sedentary behaviour. Brit J Sport Med 2020; 54: 1451–1462.

